# Assessing the efficacy of interventions to control indoor SARS-Cov-2 transmission: an agent-based modeling approach

**DOI:** 10.1101/2021.01.21.21250240

**Authors:** Trevor S. Farthing, Cristina Lanzas

**Author notes:** Correspondence to Cristina Lanzas.

## Abstract

Intervention strategies for minimizing indoor SARS-CoV-2 transmission are often based on anecdotal evidence because there is little evidence-based research to support them. We developed a spatially-explicit agent-based model for simulating indoor respiratory pathogen transmission, and used it to compare effects of four interventions on reducing individual-level SARS-CoV-2 transmission risk by simulating a well-known case study. We found that imposing movement restrictions and efficacious mask usage appear to have the greatest effects on reducing infection risk, but multiple concurrent interventions are required to minimize the proportion of susceptible individuals infected. Social distancing had little effect on reducing transmission if individuals move during the gathering. Furthermore, our results suggest that there is potential for ventilation airflow to expose susceptible people to aerosolized pathogens even if they are relatively far from infectious individuals. Maximizing rates of aerosol removal is the key to successful transmission-risk reduction when using ventilation systems as intervention tools.

**Article Summary Line:** Imposing mask usage requirements, group size restrictions, duration limits, and social distancing policies can have additive, and in some cases multiplicative protective effects on SARS-CoV-2 infection risk during indoor events.

## 1. Introduction

Understanding transmission mechanisms is necessary to generate evidence-based guidance for controlling infectious diseases. Severe Acute Respiratory Syndrome Coronavirus 2 (SARS-CoV-2), the causative agent of Coronavirus Disease 2019 (COVID-19), is primarily spread through infectious respiratory droplets and aerosols of varying size (*1, 2, 3, 4, 5*). These media are expelled when an individual speaks, coughs, sneezes, or otherwise expectorates (*6, 7*). Pathogen transmission can occur when these virion-containing particles are inhaled by, or otherwise come into contact with the mucosae or conjunctiva (i.e., mouth, nasal membranes, or eyes) of a susceptible person (*5*). Aerosol transmission is emerging as an important transmission pathway, particularly for large clusters associated with superspreading events (*2, 8, 9, 10*). van Doremalen et al. (2020) (*11*) found that SARS-CoV-2 can remain viable in aerosolized droplets for at least 3 hours post expectoration. While these results may not accurately represent SARS-CoV-2 stability outside of laboratory conditions (*5*), their findings are in line with case reports of viral-RNA detection in air collected from hospital rooms housing COVID-19 patients (*12, 13, 14, 15*).

Transmission of SARS-CoV-2 is more likely in indoor settings than outdoors (*8, 9*). Households are the most common venue linked to transmission, but healthcare facilities, religious venues, food processing plants or prisons are also likely to be associated with large clusters of COVID-19 cases (*9*). Recommended interventions to reduce indoor transmission include: social distancing, use of face coverings, increased ventilation, and reduced group sizes (*1*). U.S. state government recommendations and restrictions for group size limits in indoor gatherings range from 10 to 100 people or 10 – 75% of a locale’s original capacity (*16*). There is little evidence-based research to support specific group size restrictions, however, and few studies have sought to identify the most-effective strategy for limiting indoor SARS-CoV-2 transmission.

Some mathematical models have been built to support individual-level risk assessment of indoor transmission and analyze aerosol contributions to past outbreaks. Chande et al. (2020) (*17*) created a tool to assess the U.S. county level probability that someone infected with SARS-CoV-2 will attend events of varied sizes. Their tool is useful for estimating the probability that SARS-CoV-2 transmission could occur during any gathering, but provides no direct measure of transmission risk from infectious individuals during events and no way to assess the impact of intervention strategies other than reducing group sizes. Other models have sought to determine the role that aerosolized infectious droplets play in indoor SARS transmission relative to larger droplets that are unlikely to be inhaled, and quantify the transmission risk attributable to aerosols in varied environments (*18, 19, 20, 21, 22*). These models are primarily based on Wells-Riley equations for estimating aerosol-attributable risk, which assume homogenous spatiotemporal mixing of air constituents and exposure to infectious agents (*23*). Mathematical indices and parameter values in these models can be adjusted to simulate effects of intervention strategies like social distancing (*22*) and increased ventilation rates (*18, 20, 21, 22*), but are insufficient for capturing or accounting for any behavior- or environment-mediated spatiotemporal heterogeneity in transmission risk. Shao et al. (2021) (*24*) used a fluid dynamics model to simulate ventilation effects on SARS-CoV-2 transmission while allowing for heterogenous droplet movement behaviors. Their findings highlight the need to account for within-room spatial heterogeneity when studying indoor transmission risk, as phenomena like ventilation can increase infection risk to individuals in one area of a room or building while simultaneously mitigating risk in another.

Here, we present a spatially-explicit agent-based model (ABM) for simulating within-room respiratory pathogen transmission to inform policy-making decisions aiming to mitigate indoor transmission and implementing individual-level interventions. By simulating spatiotemporal droplet dynamics (e.g., emission of varying droplet size and subsequent distribution in the environment) as well as allowing for dynamic movement and positioning of infectious and susceptible individuals, our model allows virion exposure rates to vary within indoor settings. We use our model to estimate effects of proposed COVID-19 intervention strategies for indoor environments (i.e., increased airflow, limiting contact durations, wearing masks, and increased interpersonal spacing). For benchmarking purposes, we simulate the outbreak that took place during a choir practice in Skagit County, WA in March 2020 (*2*). Additionally, we further investigate potential drivers of superspreading events, like the Skagit County example, by characterizing and comparing how different aspects of indoor gatherings (i.e., population density, duration, quanta production by infectious individuals, and ventilation effects) impact transmission risk. Through these analyses we provide guidance for minimizing SARS-CoV-2 transmission during indoor gatherings.

## 2. Methods

### 2.1 Model Description

We developed a spatially-explicit, stochastic ABM to simulate both direct-droplet and airborne respiratory pathogen transmission in indoor settings. This model was created and executed using the open-source modeling software, NetLogo (Ver. 6. 1. 1) (*25*) and is available for download at https://github.com/lanzaslab/droplet-ABM. In Appendix S1 we provide a detailed description of our model in accordance with ODD (Overview, Design concepts, Details) standards outlined by (*26*). We present a limited summary of the model design below. When describing infectious media in our model, we use the term “droplet” to refer to respiratory droplets of any size.

Agents in our model represent people congregating in a fixed space (e.g., students in a classroom, diners in a restaurant, etc.). Patches (i.e., grid cells in the NetLogo model interface) represent 1 x 1 m^2^ areas, and the spatial extent can range from 1 to ∞ m^2^. The model time step is 1 minute. Droplets ranging from 3 to 750 *μ*m in diameter are expelled by infectious agents. Subsequently, droplets can be inhaled, fall out, diffuse to nearby patches, move via directed airflow, and decay at fixed rates over the course of a simulation. Infection in our model is driven by exposure to virions contained in these droplets, and the number of virions per droplet scales with droplet size. The rate at which droplets fall out (i.e., are removed from circulating air flows) of the simulation is based on the calculated terminal velocity falling speed for droplets, and therefore varies with droplet size. Droplet sizes incapable of settling on the ground within one minute are allowed to move between patches via ventilation- and diffusion-induced airflow. Thus, risk of exposure and subsequent infection for susceptible individuals varies by space and time during the simulation (Figure 1). We recognize that the ability of forced air ventilation systems to reduce local respiratory pathogen transmission is linked to their ability to move aerosolized droplets away from susceptible individuals in three dimensions (*18, 24*). Though this effect is not explicitly tied to airflow inputs in our model, which only allows airflow in two dimensions, we can effectively simulate ventilation-induced aerosolized droplet movement to heights outside of individuals’ inhalation ranges by increasing the decay rate when ventilation effects are simulated. In addition to controlling the number of individuals present and the size of the simulated world, users can dictate infectiousness parameters and other scenario-specific variable values (e.g., number of infectious individuals, probability that infectious individuals are asymptomatic, cough frequency, number virions per mL of droplet fluid, risk of infection given exposure to 1 virion, etc.), ventilation parameters (e.g., direction and speed of airflow, droplet filtration probability, etc.), and adherence to transmission-risk-reduction guidelines (e.g., mask usage, local social distancing, etc.).

**Figure 1.**
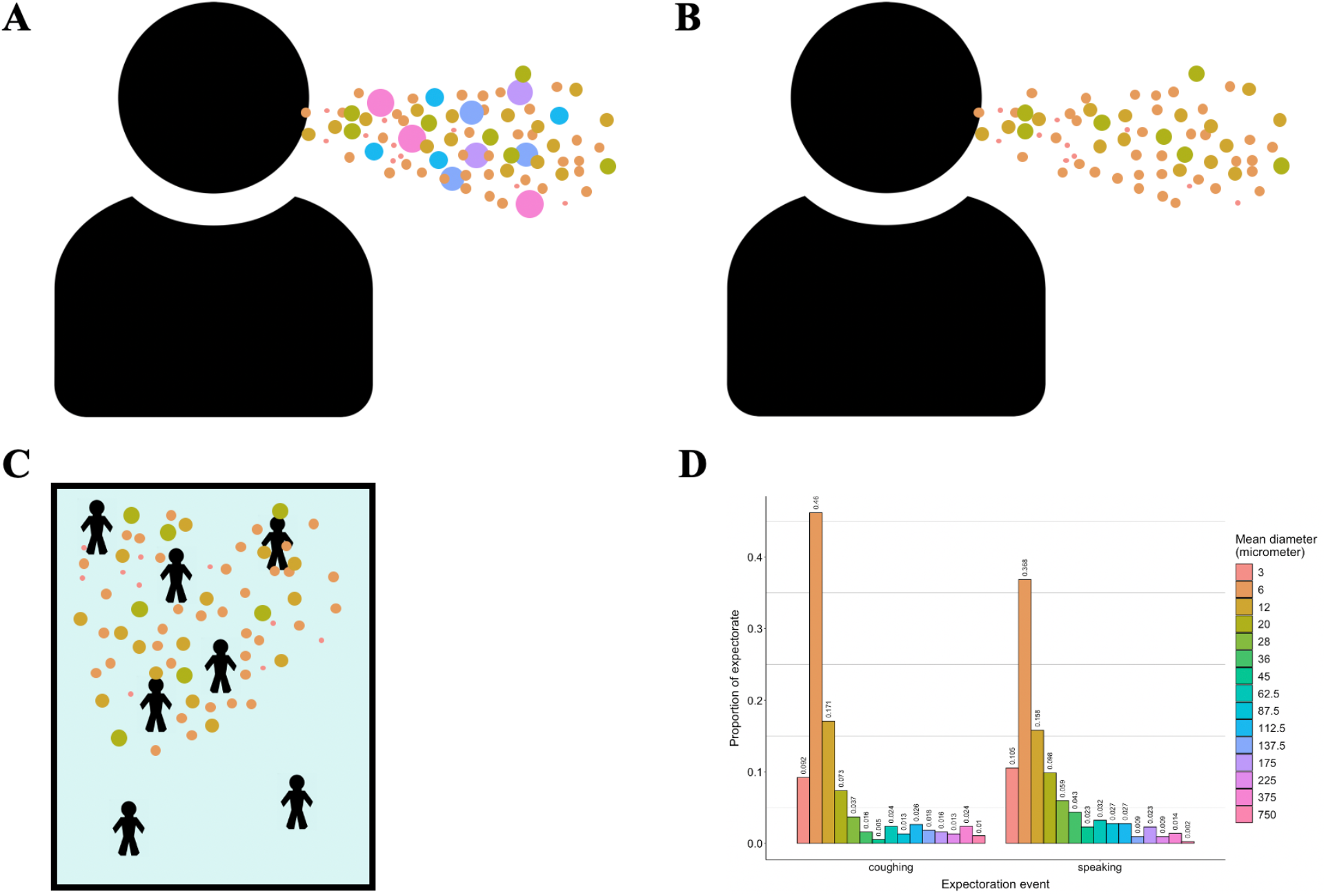
Model droplet dynamics. A) Infectious individuals expel droplets of different sizes. B) Relatively large droplets fall out of the air quickly post expectoration. C) Smaller droplets remain aerosolized for longer time periods and move throughout the simulated room via diffusion and forced airflow effects. D) Distribution of droplet sizes during expectoration events. Distributions of size classes during coughing and speaking events are based on findings of (*47*), and represent mean observed droplet-size measurements they recorded 60 mm away from individuals’ mouths immediately following these activities.

### 2.2 Testing SARS-CoV-2 transmission reduction strategies

#### 2.2.1 Case scenario and model inputs

In March 2020, there was a probable SARS-CoV-2 superspreading event during a choir practice taking place at a church in Skagit County, Washington, USA (*2*). Sixty-one people were in attendance, one attendee was experiencing flu-like symptoms at the time and later tested positive for COVID-19 (*2*). This individual likely infected 53 other attendees over the course of the event (*2*). We briefly describe our rationale for setting scenario-specific input values to simulate this case in our model below, but more-detailed explanations for input and parameter values are given in Appendix S2, and Appendix S3 describes how sensitive simulated infection risk is to variations in select model parameters.

We know from (*2*) that the choir practice lasted 150 minutes in total, split into 4 distinct time intervals. In our simulations, we decided to rearrange agents in our model after 40, 90, and 105 minutes to recreate mixing associated with changing time intervals. At timestep 105, individuals moved back to their initial placements, representing their adherence to assigned seating during interval 4 (i.e., minutes 105 – 150). The seating chart has not been shared due to privacy concerns (*21*) however, we can assume that a maximum of 2 people could be within 1-m^2^ patches in this scenario. We set the inhalation rate for simulated individuals to 0.023 m^3^ air/min, a rate consistent with adults participating in light activity (*27*). Because it is uncertain whether or not the forced-air system was turned on during the choir practice (*21*), we decided to run our simulations in two sets: ventilation-on (i.e., both forced-air effects and natural diffusion moved droplets between patches) and ventilation-off (i.e., only natural diffusion moved droplets between patches).

In addition to model the baseline scenario, we modulated values of model inputs related to group-level risk-reduction strategies (i.e., limited population, limited contact durations, mask usage, and meter-level social distancing) between simulations in order to assess the efficacy of each strategy on reducing the number of susceptible individuals infected. Regarding mask usage, we assumed face coverings have both source-prevention and wearer-protection effects, and reduced global droplet exposure/exhalation rates by 0%, 25%, 50%, 75% and 90%. The upper range here is intended to simulate the use of N95 and simple surgical masks, which are estimated to reduce aerosol emission rates by approximately 90% and 74%, respectively (*28, 29*). Lower values are intended to simulate the use of single- and multi-layered fabric masks, for which a wide range of aerosol-filtration efficacies have been reported (*30*). When simulating mask usage, we assumed that all individuals were wearing masks and that all masks had the same efficacy. Table 1 outlines the model input values for our superspreading-scenario simulations.

**Table 1.**
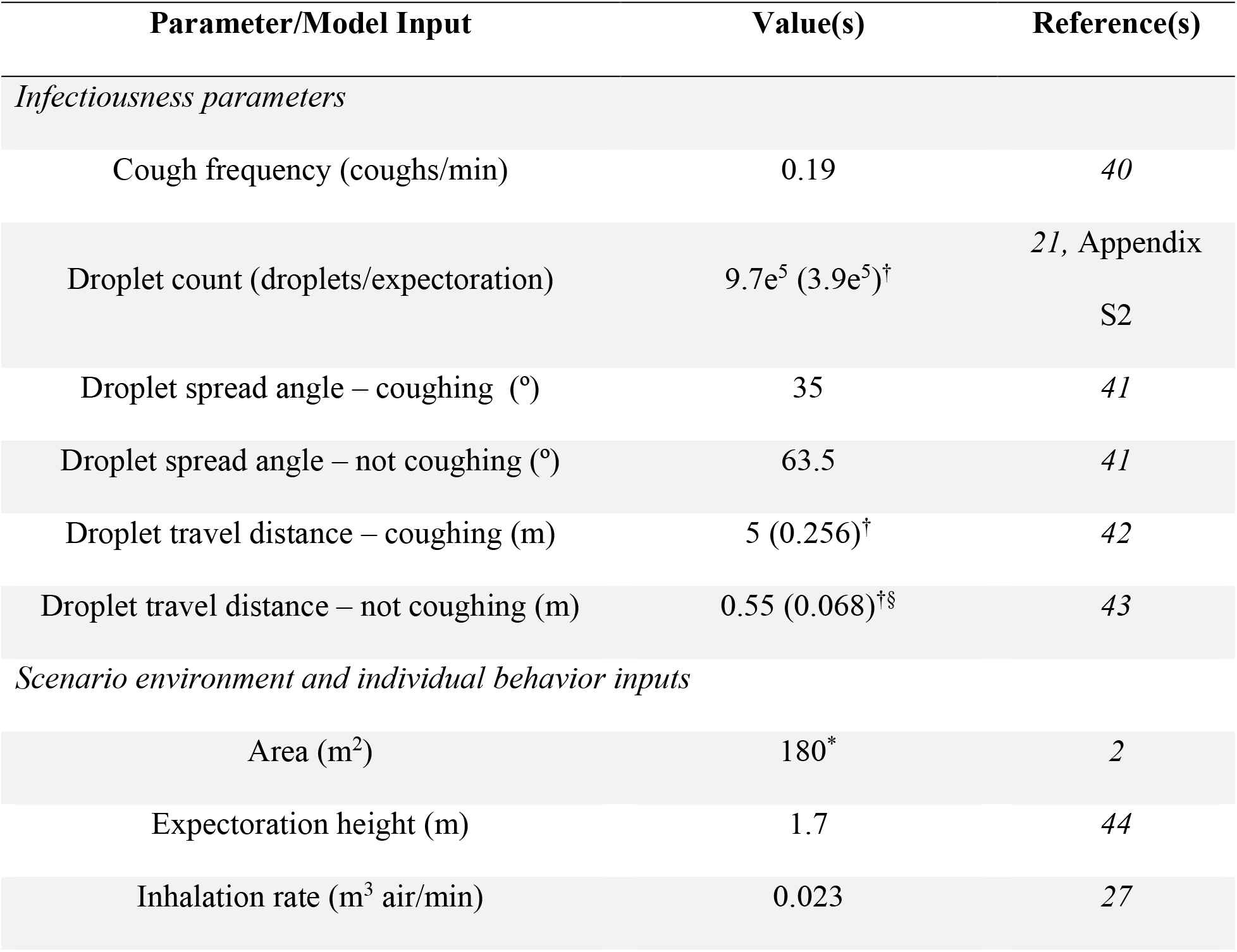
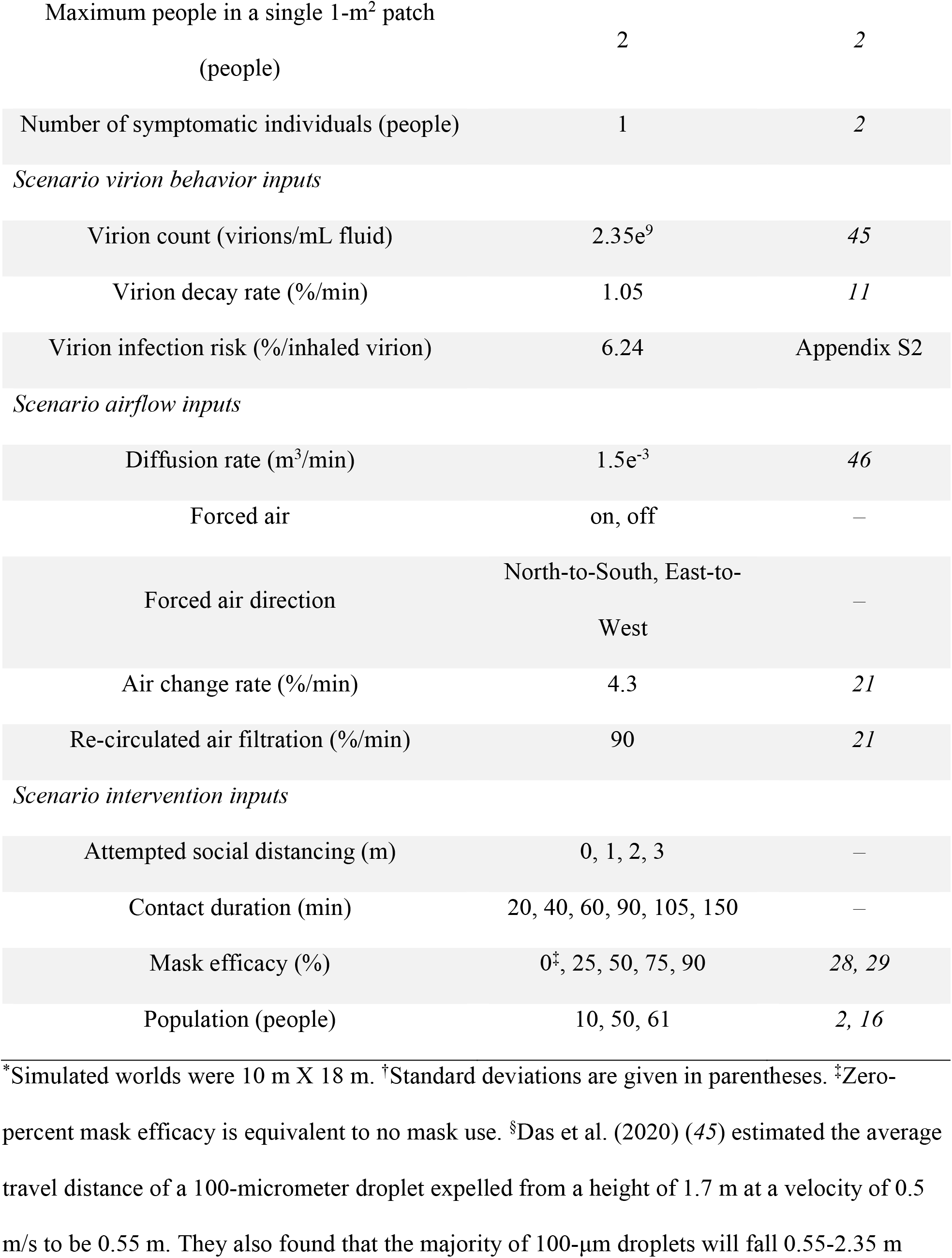
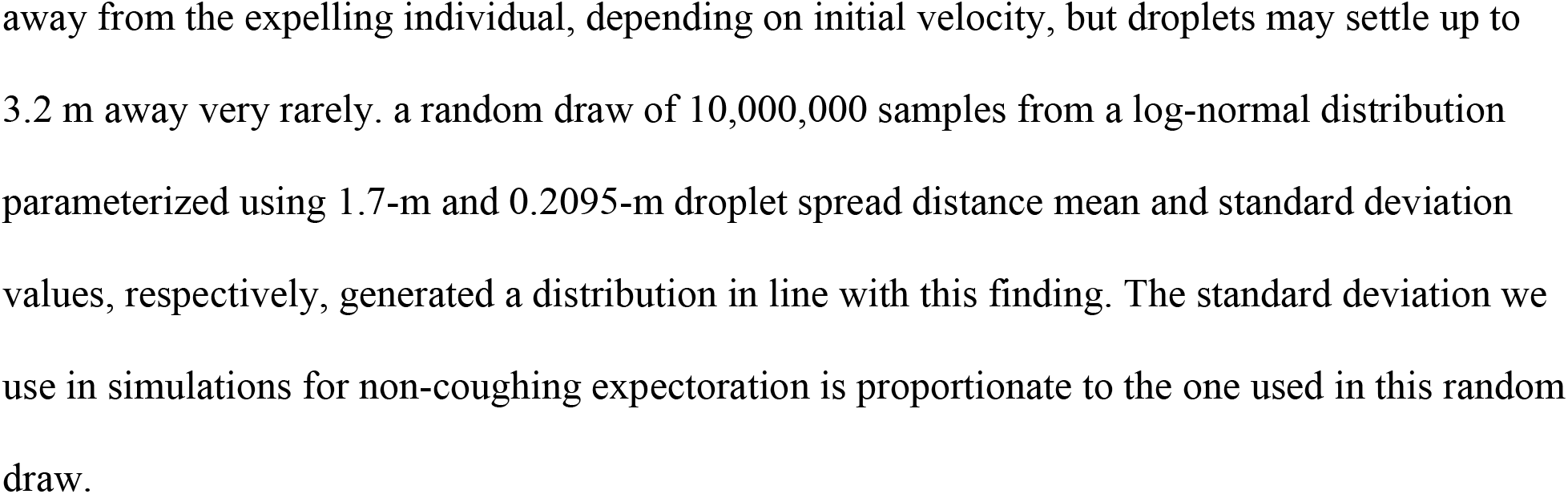
Parameter descriptions for simulations of the Skagit County, WA March 2020 SARS-CoV-2 transmission case study.

#### 2.2.2 Running simulations and analyses

We set up a factorial simulation run within the NetLogo BehaviorSpace using our specified input levels. We ran 1000 replicates of all input set combinations, ultimately resulting in 1,080,000 simulations. Simulations were aggregated into a single data set prior to analysis.

We used a beta regression model with a fixed unknown precision parameter, *ϕ*, (*31*) to estimate effects of interventions on the mean proportion of susceptible individuals infected in our full simulation set, *μ*. Beta regression models are employed to analyze proportion data (*31, 32*). We chose to use this method because the potential number of infected individuals in each simulation was limited by the simulated group size, which was a predictor variable of interest, and because preliminary analysis suggested that fitting our data to a beta distribution better explained observed variation than other regression models. We therefore determined it was more appropriate to evaluate effect sizes in terms of the relative proportion of susceptible people infected rather than their total number. We fit our data to the model: 

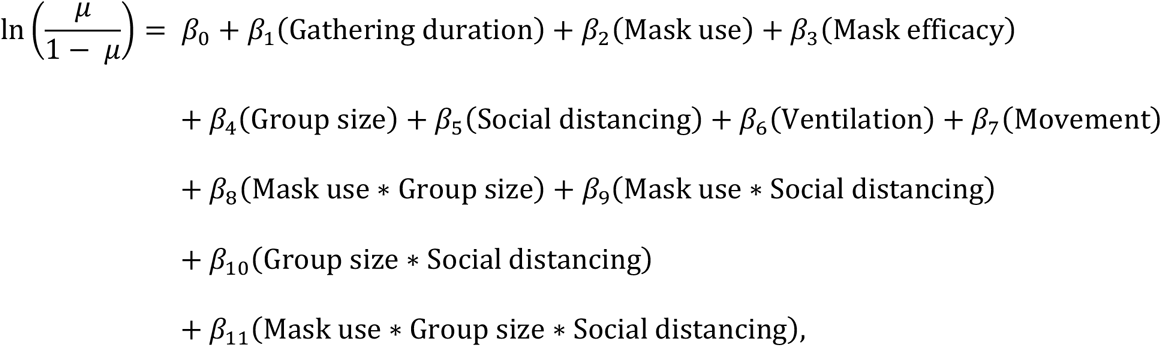

where “Gathering duration,” “Mask efficacy,” and “Group size” are intervention-strategy variables relating to: minutes of simulated interaction between individuals, the efficacy of worn face masks for reducing expectoration and inhalation of infectious droplets, the simulated population size, and attempted meter-level social distancing in each realized simulation, respectively. The variables “Mask use,” “Movement,” and “Ventilation” are known confounders related to: reduced droplet spread distance from expectorating infectious individuals wearing masks, the number of times individuals were rearranged within simulations to reflect mixing of event attendees, and movement of infectious aerosols throughout the simulated space due to a forced-air ventilation system, respectively. “Mask use” is a binary variable taking the value of 1 when simulated individuals are masked (i.e., “Mask efficacy” > 0), and 0 when they are not. “Movement” takes any one value within the range of 1-to-4, dependent on “Gathering duration.” “Ventilation” is a binary variable taking the value of 1 for simulations in the “ventilation-on” subset, and 0 for those in the “ventilation-off” subset.

Because beta regression procedures assume all dependent variable values fall between 0 and 1, we used the data transformation procedure described by (*33*) to reconstruct our proportion data without these extremities. All analysis and plotting was carried out using functions from the “betareg” R package (*32*) in RStudio (v. 1.1.463) (*34*) running R (v. 3.6.2) (*35*). We calculated a pseudo-R^2^ (*31*) to assess goodness of fit for our regression model

### 2.3 Evaluating drivers of transmission in indoor gatherings

To assess the relative contribution of environmental conditions to SARS-CoV-2 transmission risk, we conducted a sensitivity analysis to ascertain relative effects of population density, gathering duration, quanta production by infectious individuals, and ventilation on SARS-CoV-2 infection risk beyond the conditions tested in the Skagit County case. In addition, we quantified the ability of different ventilation system aspects (i.e., air-change rate, filtration rate, and effective three-dimensional droplet removal rate) to reduce SARS-CoV-2 transmission risk. Table 2 describes the model input values for these indoor-gathering-risk-assessment simulations.

**Table 2.**
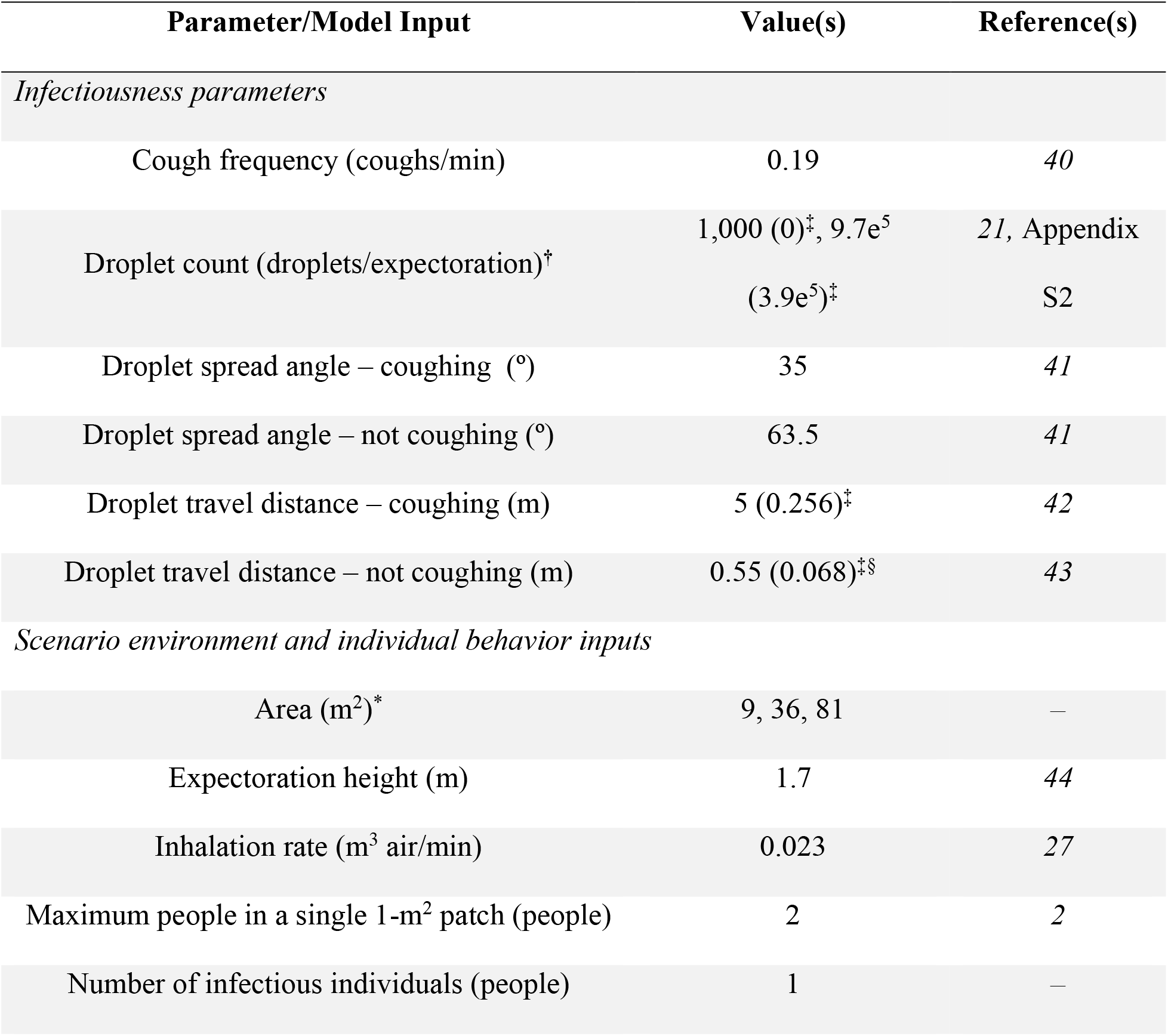
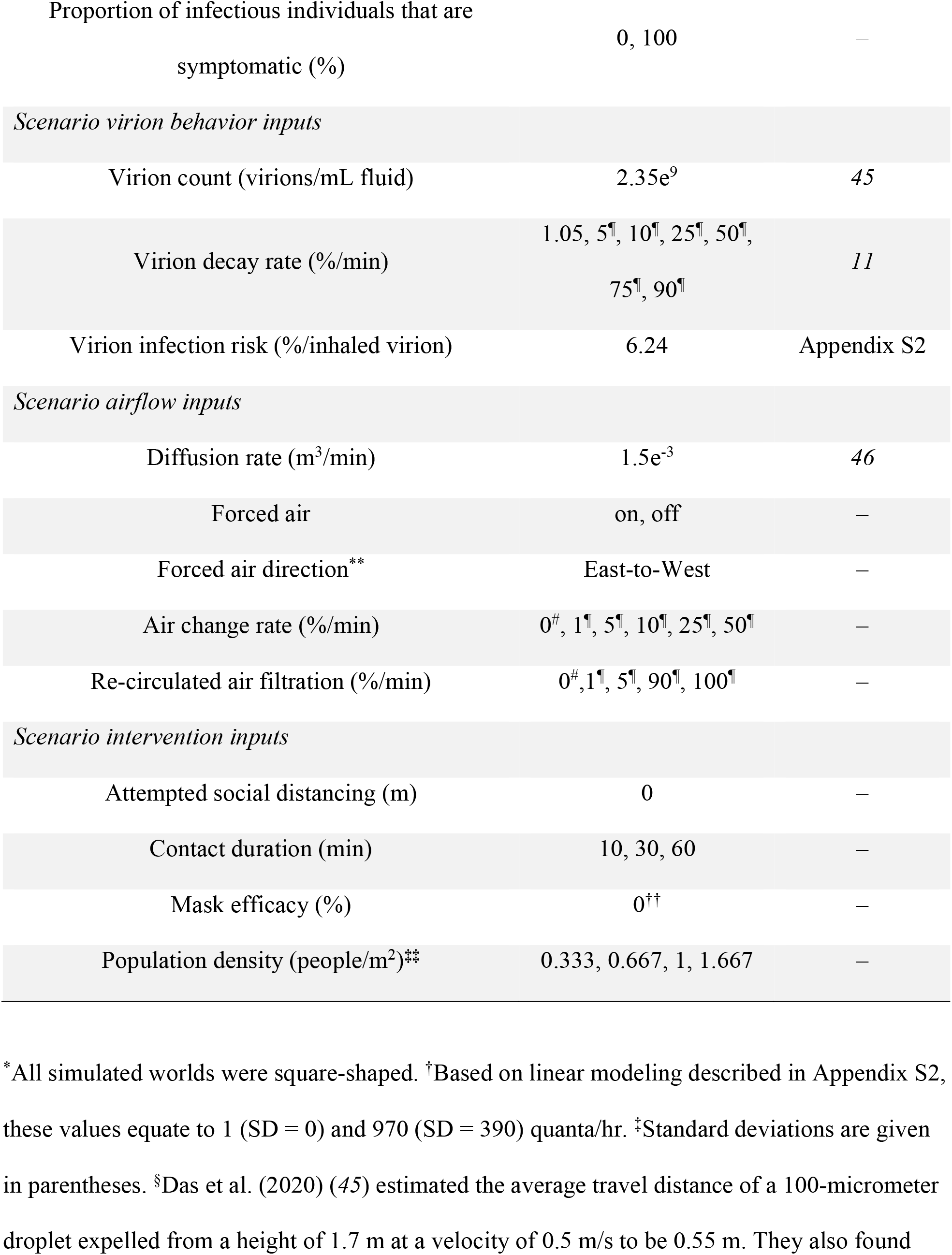
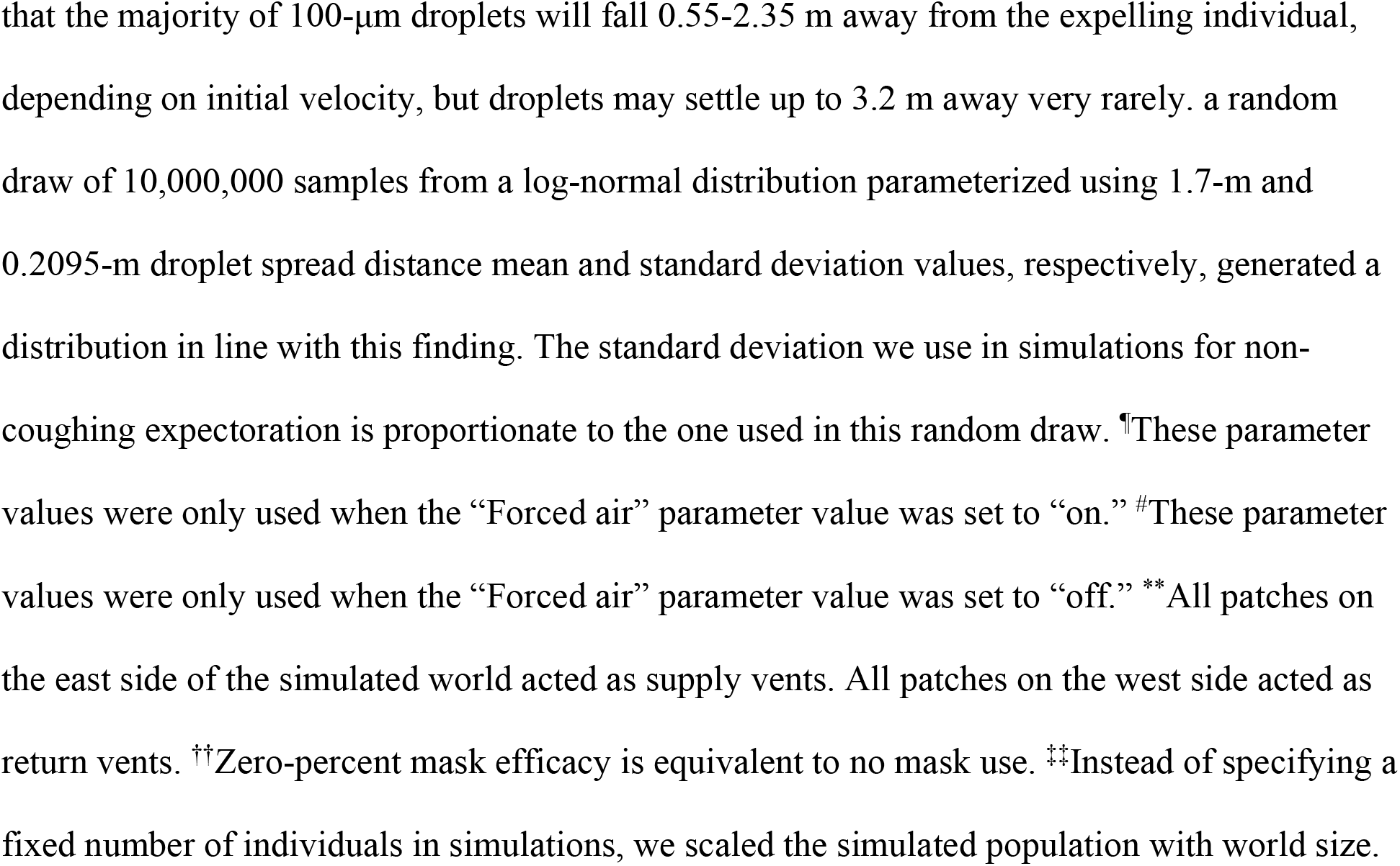
Parameter descriptions for ventilation-system effect evaluations.

We set up a factorial simulation run within the NetLogo BehaviorSpace using our specified input levels. We ran 1000 replicates of each parameter set combination when the “Forced air” parameter was set to “on” and when it was “off.” We ran these sets separately in order to save computation time as there were many inputs that only changed when forced airflow was simulated. Ultimately, we produced 144,000 “off” simulations, and 20,160,000 “on” simulations. In both sets, we identified simulations when transmission occurred (i.e., simulations where ≥ 1 person was infected), and recorded this occurrence as the binary variable *y*_*i*_ so that 

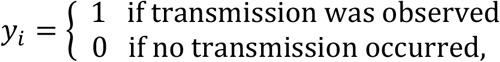

for each realized simulation, *i*.

We aggregated simulation data into a single data set and carried out a logistic regression analysis to estimate effects of variable inputs on observed differences in the probability of observing ≥ 1 infections. We fit our data to the model: 

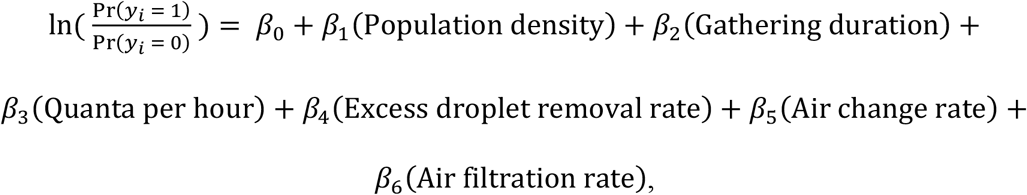

where “Population density” is given by 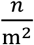, and the “Excess droplet removal rate” (%/min) represents the increased removal of aerosols due to ventilation-induced 3-dimensional droplet movement. It is given by the equation: (Virion decay rate − 1.05). The 1.05 here represents the general SARS-CoV-2 decay rate (i.e., 1.05%/min) as reported by (*11*). Subtracting this baseline value from the simulated Virion decay rate gives us an excess removal rate that we use as a proxy for 3-dimensional droplet removal attributable to forced airflow movement. When no forced airflow is simulated, excess droplet removal, air change, and filtration rates all equal 0. We calculated the Tjur (*36*) pseudo-R^2^ for our logistic regression model to assess goodness of fit.

## 3. Results & Discussion

We presented a stochastic ABM for studying indoor individual-level respiratory pathogen transmission, and used it to demonstrate the potential effectiveness of multiple intervention strategies for reducing SARS-CoV-2 transmission in an indoor group setting mimicking that of a known superspreading event. We were able to effectively recreate the empirical proportion of susceptible individuals likely infected during the Skagit County superspreading event by simulating the gathering without implementing any intervention strategies (Figure 2).

**Figure 2.**
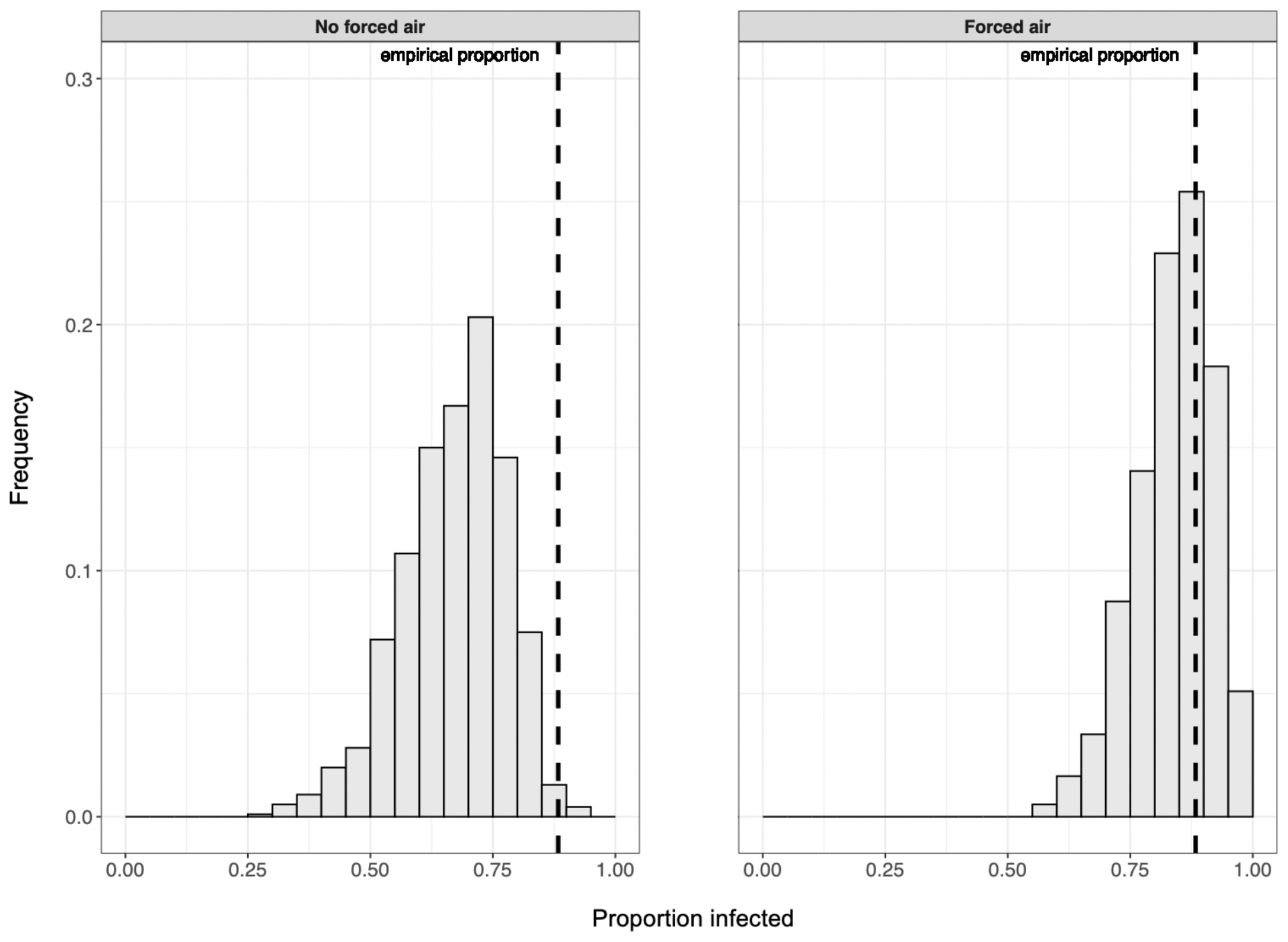
In the absence of interventions to reduce transmission risk, the proportion of susceptible people infected in simulations can reflect the case study value (i.e., 0.88) and is more likely to do so when forced airflow is included.

Our beta regression model for estimating intervention efficacy had a pseudo-R^2^ of ≈ 0.43. Given the number of stochastic processes in our ABM, the explanatory power of the model is acceptable. Duration limits and efficacious mask usage appear to have the greatest effects on reducing the proportion of susceptible individuals infected, but multiple concurrent interventions are required to minimize the proportion of susceptible individuals infected (Table 3, Figure 3). However, it is important to note that observed proportional differences are more meaningful for relatively large groups than for smaller ones. The effectiveness of limiting the duration of gatherings for reducing the proportion of infected individuals appears to largely result from reducing the confounding movement effect that increases over time, thereby reducing the probability that susceptible individuals will move from uncontaminated space to areas with greater concentrations of infectious aerosols or nearby to infectious individuals where they may be exposed to large virion-containing droplets (Table 3). We show that simply by limiting the time spent rehearsing that night to 40 min, reducing random mixing between attendees by ending the event prior to splitting into disparate groups (*2*), the proportion of people infected could have been reduced by 70 – 88% even without implementing any other intervention strategies. Therefore, imposing movement restrictions could be a more effective intervention than strict duration limits.

**Table 3.** Logit scale estimates associated with 1-unit increases in covariate values given by our beta-regression model for evaluating intervention effects. Wald 95% confidence intervals are given in parentheses.

| Coefficient | Estimate | *p* |
| --- | --- | --- |
| **Intercept** | -2.927 (-2.940, -2.914) | — |
| $\phi$ | 5.808 (5.791, 5.824) | — |
| **Gathering duration (min)** | 0.012 (0.012, 0.012) | < 0.001 |
| **Mask efficacy (%)** | -0.015 (-0.015, -0.015) | < 0.001 |
| **Mask use** | -0.949 (-0.964, -0.935) | < 0.001 |
| **Movement (No. rearrangements)** | 0.491 (0.485, 0.497) | < 0.001 |
| **Group size (people)** | 0.001 (0.001, 0.001) | < 0.001 |
| **Social distancing (m)** | -0.250 (-0.256, -0.243) | < 0.001 |
| **Ventilation** | 0.898 (0.895, 0.902) | < 0.001 |
| **Mask use : Group size** | 0.014 (0.013, 0.014) | < 0.001 |
| **Mask use : Social distancing** | -0.018 (-0.025, -0.010) | < 0.001 |
| **Group size : Social distancing** | 0.004 (0.004, 0.004) | < 0.001 |
| **Mask use : Group size : Social distancing** | 1.923e<sup>-4</sup> (3.039e<sup>-5</sup>, 3.542e<sup>-4</sup>) | 0.020 |

**Figure 3.**
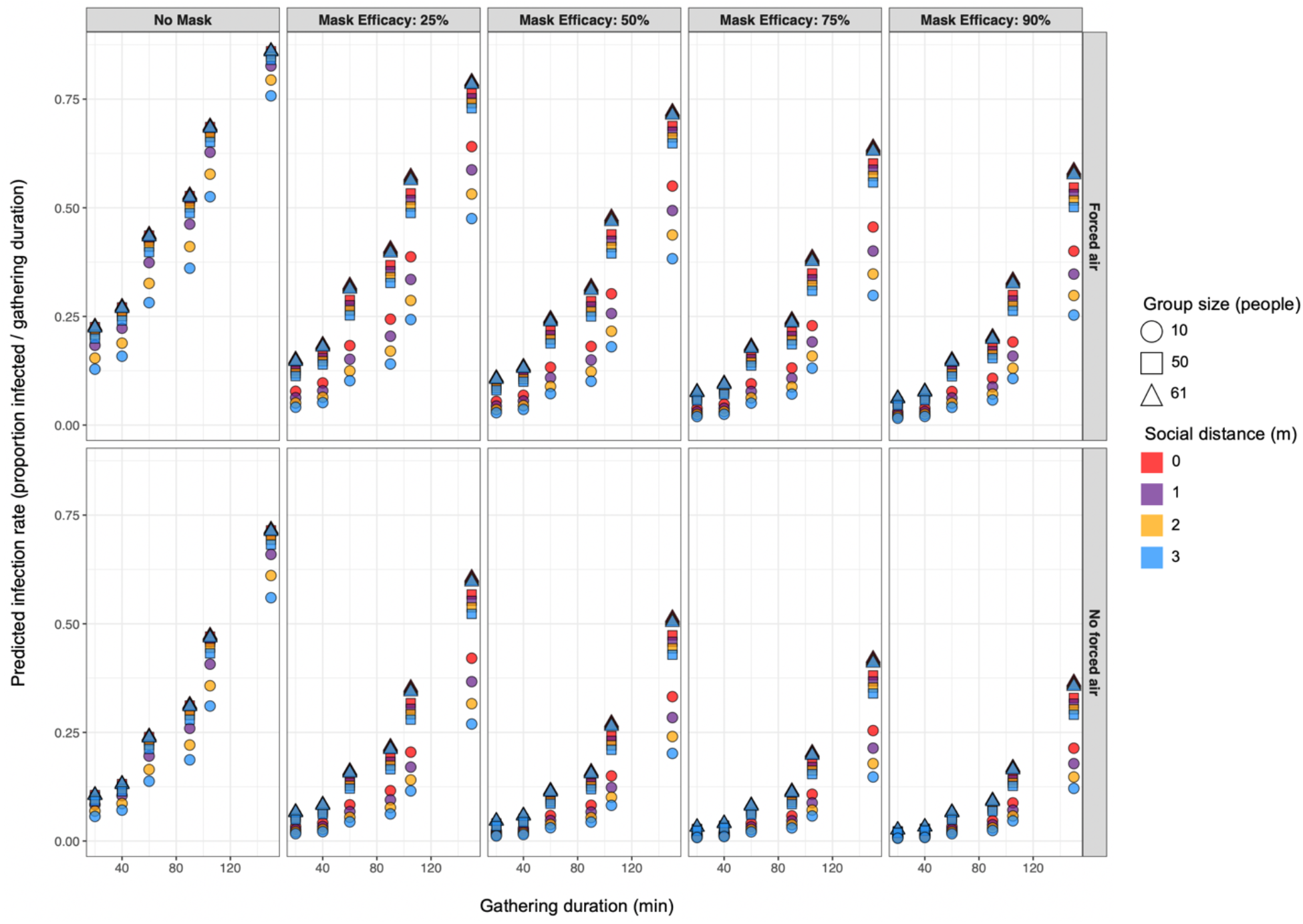
Predicted proportion of susceptible populations infected with SARS-CoV-2 for varied parameter sets suggest that concurrent deployment of multiple interventions is required to achieve near-zero transmission rates.

We found that mask usage and social distancing interventions are relatively more effective for reducing proportional infection rates in small groups than in large ones. Our findings suggest that in the Skagit County choral case, duration limits with implied movement restrictions and mask usage would have been the most-effective intervention strategies for reducing SARS-CoV- 2 infection rates, but multiple interventions would have needed to be deployed simultaneously to reach near-zero rates (i.e., mean rate < 0.5 people / gathering duration). Infection rates generally increased with group size and decreased with mask efficacy, and we found that when movement rates were minimized (i.e., gathering durations ≤ 40) we could minimize infection rates with relatively-low mask efficacy or even no mask usage in some cases. Our results support recent evidence suggesting that even wearing masks with relatively low droplet-filtering abilities around others can help to reduce exposure to infectious agents (*30, 37*). Attempted social distancing up to 3 m had little effect on transmission rates relative to other intervention strategies. That said, because social distancing generally had a greater effect on proportional infection rates when group size was limited to 10 people, and 2-m social distances reduced the mean number of infections in larger groups, we can intuit that the relatively small overall effect of social distancing was likely due to the presence of physical barriers (e.g., edges of the simulated world) or the physical arrangement of nearby individuals impeding agents’ attempts to social distance, rather than due to far-reaching droplet spread that makes social distancing irrelevant.

Conclusions regarding social distancing effects are further supported by our logistic regression model results that describe the relative effects of population density, gathering duration, quanta production, and ventilation on the probability of indoor SARS-CoV-2 transmission from a single infectious individual (Table 4). This model had a pseudo-R^2^ value of 0.25 and demonstrated that among the considered variables, population density was the most-important contributor to SARS-CoV-2 transmission risk. Additionally, increases in gathering duration, infectious aerosol production, and horizontal air movement all escalate the probability that transmission will occur during gatherings, though the effect is much lesser than that of increasing population density. The relatively small effects of quanta production and duration on transmission risk suggest that once individuals are exposed to infectious agents, they are likely to become infected quickly. Thus, minimizing susceptible people’s exposure to infectious media is of paramount importance for controlling COVID-19 incidence.

**Table 4.** Logit scale estimates associated with 1-unit increases in covariate values given by our logistic-regression model for evaluating effect on SARS-CoV-2 transmission risk during an indoor gathering. Wald 95% confidence intervals are given in parentheses.

| **Coefficient** | **Estimate** | **Odds ratio** | ***p*** |
| --- | --- | --- | --- |
| **Intercept** | -0.146 (-0.151, -0.140) | — | — |
| **Population density**<br>**(people/m<sup>2</sup>)** | 2.766 (2.761, 2.771) | 15.891 (15.813, 15.968) | < 0.001 |
| **Gathering duration (min)** | 0.015 (0.015, 0.015) | 1.015 (1.015, 1.015) | < 0.001 |
| **Quanta (quanta/hr)** | 0.002 (0.002, 0.002) | 1.002 (1.002, 1.002) | < 0.001 |
| **Excess droplet removal rate**<br>**(%/min)** | -0.024 (-0.024, -0.024) | 0.976 (0.976, 0.976) | < 0.001 |
| **Air change rate (%/min)** | 0.017 (0.017, 0.017) | 1.02 (1.02, 1.02) | < 0.001 |
| **Air filtration rate (%/min)** | -0.005 (-0.005, -0.005) | 0.995 (0.995, 0.995) | < 0.001 |

Regarding observed effects of ventilation in our beta and logistic regression models, our results suggest that in spite of some evidence that forced-air ventilation systems can reduce risk of respiratory pathogen infection from indoor aerosols (*38, 39*), there is potential for forced airflow to expose susceptible people to aerosolized pathogens even if they are relatively far from infectious individuals, and therefore increase transmission risk. We show that, though filtering re-circulated air can lower transmission risk (Table 4), increasing this effect is unlikely to compensate for the elevated risk attributable to increased horizontal air-change rates (Tables 3 & 4). It appears that maximizing rates of three-dimensional aerosol removal is the key to successful transmission-risk reduction when using forced-air ventilation systems as intervention tools. Our results are therefore consistent with the findings of (*18*), who advise that “displacement” ventilation systems, those designed to vertically stratify indoor air by temperature and remove warmer air, are likely able to reduce local SARS-CoV-2 transmission risk. “Mixing” ventilation systems, designed to distribute temperature and aerosols equally throughout the space, are likely insufficient for preventing transmission and may even facilitate it (*18*).

Given our findings, we maintain that in areas where COVID-19 prevalence remains high, holding events associated with relatively-increased mixing rates between individuals (e.g., social gatherings, sporting events, etc.) should be avoided even if attendance rates are presumed to be low. Such events are likely to be associated with SARS-CoV-2 transmission if ≥ 1 infectious individual(s) were to attend, the probability of which increases linearly with group size (*17*). It is important to note however, that though our results provide insight into mechanisms for reducing SARS-COV-2 transmission rates, given the effect that model parameters can have on simulation outcomes (*see* Appendix S3), our findings may not be reasonably extrapolated to accurately predict transmission in scenarios dissimilar from those we modeled here (e.g., ≥ 2 infectious individuals, fewer aerosolized virions produced during expectorations, etc.). Regardless, we can still conclude that imposing mask usage requirements, group size restrictions, duration limits, and social distancing policies have additive, and in some cases multiplicative protective effects on individual-level SARS-CoV-2 infection risk during gatherings, and can be particularly efficacious interventions when deployed simultaneously.

## Data Availability

The model referred to in our manuscript is available for download at https://github.com/lanzaslab/droplet-ABM.

## Acknowledgments

This work was partially supported by CDC U01CK000587-01M001 and R35GM134934.

## Author Bio

Trevor S. Farthing is a PhD candidate in the Comparative Biomedical Sciences program at North Carolina State University. His work combines ecological and epidemiological analyses with agent-based infectious disease modeling to quantify and compare driving forces of pathogen transmission in varied pathogen-host systems.

Dr. Cristina Lanzas is an Associate Professor of Infectious Disease at North Carolina State University. Her research focuses on the epidemiology and ecology of infectious diseases in animal and human populations. Her work combines data science, epidemiological analysis and mathematical models to study transmission mechanisms, and to identify and design effective control measures to reduce the public health burden associated with infectious diseases.

