## Appendix S1 for "Assessing the efficacy of interventions to control indoor SARS-Cov-2 transmission: an agent-based modeling approach"

### Appendix S1 – ODD Model Description

We developed a spatially-explicit, stochastic agent-based model (ABM) to simulate airborne and direct droplet-mediated respiratory pathogen transmission in indoor settings. This model was created and executed using the open-source modeling software, NetLogo (Ver. 6. 1. 1 – Willensky 1999). Below, we provide a detailed description of our model in accordance with ODD (Overview, Design concepts, Details) standards outlined by Grimm et al. (2020).

#### 1. *Purpose and patterns*

The purpose of this model is to quantify the effect of increasing group density on the probability of respiratory pathogen transmission from infectious individuals to susceptible ones, given varied spatial dimension and risk-reduction behavior (e.g., mask use, social distancing, etc.) levels in indoor settings. The ability of our model to accurately simulate infection events is predicated on its ability to recreate four processes involved in transmission: 1.) Susceptible individuals become infected through inhalation of virions contained within infectious droplets of varying sizes. 2.) Infectious agents expel infectious droplets of varying sizes, and droplets' movement, fallout, and virion-carriage rates vary with droplet size. 3.) Symptomatic infectious agents are likely to infect more susceptible individuals than asymptomatic ones, as coughing expels infectious droplets farther than does breathing or speaking alone (Kwon et al. 2012). 4.) Susceptible individuals' probability of infection can be lessened if individuals employ extra measures to avoid transmission (e.g., wearing face masks).

#### 2. *Entities, state variables, and scales*

There are two mobile agents (i.e., NetLogo agents capable of movement) in our model: *People* and *AirArrows*. *People* in our model represent people congregating in fixed space (e.g., students in a classroom, people watching a movie in a theatre, etc.), while *AirArrows* control the direction of simulated airflow in the space when ventilation-induced airflow is being simulated. Patches (i.e., grid cells in the NetLogo model interface) in our model represent 1 x 1 $m^2$ areas. When a simulation begins, spawned people can be susceptible, infectious and symptomatic (i.e., these agents represent individuals who spread the pathogen via coughing, sneezing, etc.), or infectious and asymptomatic (i.e., these agents represent individuals who spread the infection through breathing or speaking alone). Over the course of any simulation, susceptible agents may become infected with a pathogen following exposure to infectious droplets expelled from symptomatic and/or asymptomatic agents. The global environment dictates the size of the fixed space, total number of agents in the model, number of these agents that are infectious, as well as the dynamics and probabilities of infection events and airborne droplet movement within the model. Global, agent, and patch variables are described in Table S1-1, where we also make the distinction between parameters (i.e., static variables that are unchanging within simulations) and dynamic variables that may vary within simulations. The spatial extent of our model can range from 1 to ∞ $m^2$, and is controlled by the *grid_height* and *grid_length* parameters. Each tick (i.e., one-unit time step) in our model represents a one-minute progression.

3. *Process overview and scheduling*

Model processes are outlined in Figure S1-1 and described in detail herein. Upon initialization, a world with $grid\_height * grid\_length$ patches is generated. Following world creation, *n* susceptible people spawn within patches. People spawn one at a time and, if the *social-distance* parameter is > 0 m, they appear at a random location at least *social-distance* from any other person. If there is no available space ≥ *social-distance* from any person, newly spawned people will be placed at a random location as far away from others as possible. If *social-distance* equals 0 m, people spawn in completely random locations. Once spawning is completed, *n_infectious* people are randomly selected from the pool of susceptible agents to transition to the "infectious" health state. Infectious people have a *symp-pr* probability of being classified as "symptomatic" and a 1 - *symp-pr* probability of being "asymptomatic." All people are asked to set their heading (i.e., direction they are facing) to a random direction between 0º – 360º if *face-northward* is FALSE, or between 315º – 405º if *face-northward* is TRUE. People have a *mod-proportion* probability to wear masks in the simulation, and the affected groups (i.e., susceptible only, infectious only, or both susceptible and infectious individuals) that may transition to wearing masks are designated by the *mod_group* parameter. Masked people are then asked to update their *exposureRisk* and *expectorateRisk* variable values. The default values for each variable is 100%, but will change to the *maskRisk-mod* parameter value.

The final aspect of simulation setup is to establish agents for simulating ventilation airflow if *ventilation* is TRUE. To create a supply and return vent(s), *numSupplyVents* and *numReturnVents* patches on *ventilSupplyWall* and *ventilReturnWall* world borders are designated as supply and return vents, respectively. All non-return-vent patches are asked to spawn a single airArrow. All airArrows are asked to set their heading towards the closest return vent patch. AirArrow headings will be used to direct ventilation airflow. This concludes the simulation setup procedure.

Following setup, the simulation begins in earnest. If *ventilation* is TRUE, the first task to take place each tick is to move droplets towards return vents. We assume very simple ventilation-induced air movement within an enclosed room where air moves only towards the return vent(s), and return vent patches transfer a proportion of droplets present there to supply vent patches while also removing some droplets from the simulation. To achieve this, we ask all patches to count the number of droplets in each size class (Figure S1-2) that will be transferred to the next patch (i.e., $num_{d_p} * ventil\_movementRate$, where $num_{d_p}$ is the number of droplets of a given size class $d$ in patch $p$). Then we simultaneously ask non-return vent patches to transfer these droplets to patches 1-patch ahead of airArrows, and ask return vent patches to transfer

$$num_{d_p} * ventil\_movementRate * (1 - ventil\_removalRate)$$

droplets to supply vents, and to remove

$$num_{d_p} * ventil\_movementRate * ventil\_removalRate$$

from the simulation.

Non-ventilation related droplet removal is the second action to occur every tick. This action represents droplet/virion removal from the local environment due to inhalation by individuals, gravitational settling, and general droplet decay. For each droplet size class, we ask all patches to remove

$$\left\{\left(\frac{vol\_B}{1\text{ m} * 1\text{ m} * expectorateHeight} * People_p\right) + \left(\frac{Vt\_diam_d}{expectorateHeight}\right) + dropletDecay)\right\} * num_{d_p}$$

droplets, where $\frac{vol\_B}{1\text{ m} * 1\text{ m} * expectorateHeight}$ is the proportion of air within the patch inhaled by a single person each minute, $People_p$ is the number of people on patch $p$, $Vt\_diam_d$ is the calculated terminal velocity (i.e., the maximum free-falling speed in m/min, assuming the force of gravity acting on an object is 9.8 m/s$^2$) of droplets in a size class $d$ (Figure S1-3), and $num_{d_p}$ is the number of droplets of a given size class $d$ in patch $p$. If patches would remove > 100 % of any size class, we ask them to instead set that $num_{d_p}$ to zero. Thus, we ensure that no patch can ever have a negative number of droplets.

Next, we ask patches to diffuse droplets of all size classes to neighbors (i.e., all patches touching them) at rate *diffusionRate*. We once again ask all patches to count the number of droplets in each size class that will be transferred to the neighbors (i.e., $num_{d_p}$ * *diffusionRate*). Then, we ask patches to evenly distribute these droplets to neighbors.

Infectious people, $i \in I$, expectorate droplets and exposed susceptible individuals, $s \in S$, may become infected. Each tick, every symptomatic person has a $cough\_frequency$ probability to cough (i.e., expel droplets relatively far out from themselves), and $(1 - cough\_frequency)$ probability to expectorate in accordance with the “non-coughing” schema (i.e., expel droplets relatively close to themselves). Asymptomatic people have no chance to cough, and will expectorate in accordance with the “non-coughing” schema with 100% probability. While droplet spread distance and angle for this schema can be modulated via model input values to reflect numerous activities (e.g., speaking, breathing, etc.), the droplet size distribution is assumed to reflect that of speaking events (Figure S1-2). Thus, parameters referring to aspects of the “non-coughing” schema are coded as “speak” parameters (e.g., *speak_airflow_angle*).

During expulsion events, droplets spread to patches in front of coughing and speaking infectious people in cones with semi-vertex angles of *cough_airflow_angle* and *speak_airflow_angle*, respectively, and lengths randomly drawn from lognormal distributions. Lognormal distributions were obtained by exponentiating Poisson distributions with known means and standard deviations, in accordance with methods described by Railsback & Grimm (2011). In our model, lognormal distributions to inform droplet travel distances from coughing and speaking people are generated from known mean and standard deviation pairs: *cough_spread_dist.mean*, *cough_spread_dist.sd*, and *speak_spread_dist.mean*, *speak_spread_dist.sd*, respectively. If infectious people are wearing masks, only the patch they are in is contaminated (i.e., cones of expectoration in these cases have lengths of 0).

The number of droplets that infectious people expel at time *t*, $dropletNum_{it}$, is determined by sampling from another lognormal distribution with known means of *speak_dropletNum.mean* or *cough_dropletNum.mean*, and standard deviations of *speak_dropletNum.sd* or *cough_dropletNum.sd,* depending on if people are speaking or coughing, then multiplying this samples value by $expectorateRisk_i$. We assume that people expectorate droplets of 16 size classes, with increasingly large mean diameters. We accept the size class frequency distributions for speaking and coughing events given by Chao et al. (2009) and shown in Figure S1-2, and enforce these distributions in our model. We assume that all droplets are evenly distributed between and within contaminated patches.

After infectious individuals expectorate, we assess if any susceptible individuals will transition to infected status. The number of virions (i.e., live pathogen capable of causing infection in susceptible individuals) in a patch, $virions_p$, is given by the equation

$$virions_p = \sum_{d=1}^{D}(virionsPerML * Vol_d * num_{d_p}),$$

where $Vol_d$ is the mean volume (in mL) of droplets in each size class, calculated using the equations presented by Anchordoqui & Chudnovsky (2020). The probability that a susceptible person on patch *p* is infected at any given time is

$$pr(infection)_s = virions_p * virionRisk * \frac{vol\_B}{1\text{ m} * 1\text{ m} * expectorateHeight} * exposureRisk_s$$

where, $virions_p$ is the number of virions in the patch containing the individual.

If *numCohorts* = 1, the simulation ends after *cohort_dur* ticks have elapsed. If *numCohorts* > 1, the simulation will last for $numCohorts * cohort\_dur$ ticks. In this case, every $cohort\_dur$ ticks, if *rearrange-cohort* is TRUE, all people will move to randomly-selected patches while still adhering to *social-distance* and *personPerPatch*-cap rules. People will set a new heading in accordance with *face-northward*. If *rearrange-cohort* is FALSE, all people are killed, and an equal number of people will spawn while adhering to *social-distance* and *personPerPatch*-cap rules. All people in the new cohort will be susceptible to infection (i.e., infectious people only exist in the first cohort).

4. *Design concepts*

Infection in our model is driven by inhalation of virions contained in droplets of varying sizes. Fomite-driven transmission, by design, is outside the scope of our model. Regarding aerosol transmission, for simplicity, we assume that droplets fall from *expectorateHeight* m at terminal velocity and our droplet-size distribution represents post-evaporation sizes. These assumptions are reasonable given the rapid speed at which droplets evaporate and reach terminal velocity (Noakes *et al.* 2006; Xie *et al.* 2007; Anchordoqui & Chudnovsky 2020), and allow us to discount local humidity, temperature, and micro-scale airflow effects on the spatial distribution of droplets within the model. Droplet size class terminal velocity is calculated using the equations presented by Anchordoqui & Chudnovsky (2020), and droplet sizes incapable of settling on the ground from *expectorateHeight* m within one tick (i.e., one minute) are allowed to move between patches via ventilation- and diffusion-induced airflow. As the number of virions within a patch is dependent on the number of droplets in each size class, spatial infection-risk heterogeneity is therefore a function of global airflow parameters and the placement of infectious people throughout the simulated world. For simplicity, we assume that mechanism of droplet-mediated pathogen transmission is the same (i.e., inhalation) for droplets of all size classes. We do realize, however, that in reality larger droplets are relatively less-likely to be inhaled and instead mediate transmission through contact with unprotected mucus membranes (Milton 2020).

We assume that the volume of air in each patch at any given time is *expectorateHeight* $m^3$ (i.e., $1\text{ m} * 1\text{ m} * expectorateHeight$) and that droplets are evenly distributed within patches. Thus, the per-capita number of virions that people inhale each tick is equal to

$$virions_p * \frac{vol_B}{1\text{ m} * 1\text{ m} * expectorateHeight}.$$

Wearing a mask to reduce successful pathogen transmission in our model modulates the number of droplets expelled by infectious people and the proportion of virions inhaled by susceptible individuals. Previous research has quantified the extent to which using personal protective equipment may reduce risk of infection with a respiratory pathogen (Jefferson et al. 2008), and recent work has shown that masks reduce the number of aerosols expelled by wearers (Asadi *et al.* 2020). Therefore, we chose to use masks to modify the individual-level probability that susceptible individuals will become infected given exposure to infectious droplets in their patch (i.e., *exposureRisk*), and the number of droplets individuals will expectorate on any given tick (i.e., *expectorateRisk*). In our model, mask use scales both of these variables equally. We acknowledge that making these scaling factors equivalent may be unrealistic however, and intend to make this a focus of future model improvement if and when more detailed information on mask-induced effects on pathogen transmission become available.

As previously noted, in our model we characterize infectious people as asymptomatic or symptomatic. "Symptomatic" here refers to agents representing individuals that present any respiratory-disease symptoms (e.g., coughing, sneezing, etc.). We parameterize droplet behavior for symptomatic and asymptomatic collectives separately because we expect them to drive infections through different means. For example, asymptomatic individuals will likely spread infectious droplets by simply breathing near or talking to susceptible people. Symptomatic individuals, on the other hand, may also frequently spread droplets through coughing, sneezing, or similar events. Airflow angle and velocity associated with these means of infection are substantially different (Kwon *et al.* 2012) and as such, necessitate separate parameters if both symptomatic and asymptomatic agents can exist in simulations simultaneously.

Agents in our model have extremely limited movement (i.e., unless *rearrange-cohort* is TRUE and *num-cohorts* > 1, people will be completely unmoving), but people are spawned relatively far away from one another if the *social_distance* parameter is > 0. As such, our model is best used for estimating transmission risk associated with scenarios where individuals are generally unmoving (e.g., students in a classroom). Scenarios like students watching a presentation at the front the room or patrons attending a show in a theatre can be further emulated if users so choose by setting face-northward to TRUE. Users may also simulate well-mixed population interactions by setting *rearrange-cohort* to TRUE and *num-cohorts* > 1. Activity-specific movements may modulate infection risk (e.g., doctors must get close to patients in order to physically examine them), but are outside the scope of our model.

This is a simple model with little adaptive agent behavior, three collectives for people agents (i.e., "susceptible," "infectious: asymptomatic", and "infectious: symptomatic"), and only one action that can be considered to be a direct interaction between agents. That is, when a simulation begins or cohorts are rearranged/replaced, newly-spawned people learn where previously-spawned ones exist and attempt to ensure that sufficient space exists between themselves and others in accordance with the *social_distance* parameter value. Their objective is to maintain effective social distances to minimize infection risk. Accordingly, model outputs (e.g., the number of susceptible people infected, time to first infection, and average inter-agent distance) are influenced by emergent patterns triggered by this behavior. No other examples of adaptive behavior, sensing, prediction, or learning, as defined by Grimm et al. (2020), exist.

Stochasticity is introduced to the model in four ways during simulation initialization, then is further incorporated in four actions that take place during each subsequent time step. At initialization stochasticity is introduced when: 1.) people decide their initial placement, 2.) subsets of people are randomly designated as infectious, 3.) *mod-proportion* $*$ 100% of people *exposureRisk* and *expectorateRisk* values are changed from 1 to *maskRisk-mod*, and 4.) if *ventilation* is TRUE but *equallySpaceVents* is FALSE, return and supply vent locations will be randomly decided (though these locations will still be confined to appropriate walls of the world). During each time step, stochasticity plays a role in: 5.) determining if infectious agents expel droplets, 6.) drawing droplet travel distances from lognormal distributions, 7.) drawing the number of droplets produced in expectoration events from lognormal distributions, and 8.) assessing whether exposed susceptible agents transition to "infected" status. We incorporated stochasticity into these processes to introduce plausible variation into simulations.

The key outputs of this model are: 1.) the number of successful infections (i.e., susceptible agents' health statuses changed from "healthy" to "infected") each tick, and 2.) the time of the first successful infection in the simulation. In addition to the primary outputs, our model also keeps track of the average distance (in m) between individuals, and all "infected" people record the number of droplets of each size class contained within their patch at the time of infection. This allows us to not only assess parameter effects on transmission rates, but also estimate the proportion of people infected by aerosols.

5. *Initialization*

All global parameters aside from those controlling transmission mechanics (e.g., airflow angles, mean and standard deviation travel distances, number of virions in droplets, etc.) or airflow rates influence model initialization (i.e., how many agents and patches are created, where they spawn, and what their initial state-variable values are). Model actions associated with initialization are outlined in Section 3. Herein we discuss the rationale in allowing the aforementioned parameters to vary between simulations.

The primary purpose of this model is to assess the effect of population density on transmission risk. Population density in our model, expressed in terms of people/$m^2$, is given by the equation

$$\frac{n}{grid\_height * grid\_length}.$$

We allow the size of our modeled world to vary, in addition to *n*, as there may be an interaction between world size and *social_distance* levels that may ultimately cause the observed number of infections to vary. Afterall, the maximum distance that agents can spread out from one another is limited by the space available to them.

We tried to make the model flexible enough to test multiple hypotheses about implementing risk-reducing strategies (e.g., social distancing, mask use, etc.). This is the primary impetus adding the *social_distance*, *maskRisk-mod*, and *mod-proportion* parameters. Similarly, the *vol_B* parameter exists so that we can assess how infection risk changes in response to different group activities people may be participating in that are associated with different breathing rates (e.g., choir practice vs. attending a lecture), the *virionsPerML* and *virionRisk* parameters exist to ensure that our model can be used to simulate transmission of different pathogens for which these values are known or can be estimated.

6. *Input data*

No model processes are driven by external data. No external data are imported into the model.

7. *Sub-models*

All sub-models are comprehensively described in sections 3-5 and outlined in Figure S1-1.

### 9. *Tables*

Table S1-1. Variable descriptions.

| *NAME* | TYPE | UNIT | PURPOSE |
|---|---|---|---|
| *avg.dist* | Global, dynamic | m | Tracks the average interpersonal distance at each tick. |
| *avg.PatchInfectiousness* | Global, dynamic | - | Tracks the average probability that exposure to patch virions will lead to infection. |
| *can-sprout?* | Patch, dynamic | - | Logical variable describing if patches are far enough away from those with people in them that new people can sprout a while keeping the desired social-distance value. |
| *cohort* | Global, dynamic | - | Identify what cohort is currently being simulated. |
| *cohort-dur* | Global, static | min | The number of ticks that each cohort lasts (i.e., how long each people cohort spends in the simulation). |
| *cohort-endTime* | Global, dynamic | ticks | Denotes the tick value when the current cohort should be replaced, or when the simulation will end. |
| *cohort-person* | Person, static | - | Denotes what cohort the person belongs to. |

| | | | |
|---|---|---|---|
| *cough_airflow-angle* | Global, static | degrees | Controls the angle of airflow associated with coughing events. Affects the spread of droplets during a given droplet-expulsion event originating from symptomatic individuals. |
| *cough-frequency* | Global, static | coughs / min | Probability that cones of infection stemming from symptomatic individuals will be parameterized using cough_airflow-angle, cough_spread-dist.mean, and cough_spread-dist.sd, instead of the asymptomatic counterparts. |
| *cough_spread-dist.mean* | Global, static | m | The mean distance from symptomatic infectious people that droplets may be spread when coughing. This value will be used to generate a lognormal distribution from which droplet-expulsion-spread distance will be randomly drawn when a symptomatic agent triggers a droplet-expulsion event. |
| *cough_spread-dist.sd* | Global, static | m | The standard deviation distance, given a cough_spread-dist.mean value, from symptomatic infectious people that droplets may be spread when coughing. This value will be used to generate a lognormal distribution from which droplet-expulsion-spread distance will be randomly drawn |
| *diffusionRate* | Global, static | $m^2$ / min | The rate at which droplets spread to adjacent patches. For simplicity we assume a standardized rate for all droplet sizes. |
| *droplets_size3* | Patch, dynamic | droplets | Counts the number of droplets in a size class with a mean size of 3 micrometers. |
| *droplets_size6* | Patch, dynamic | droplets | Counts the number of droplets in a size class with a mean size of 6 micrometers. |
| *droplets_size12* | Patch, dynamic | droplets | Counts the number of droplets in a size class with a mean size of 12 micrometers. |
| *droplets_size20* | Patch, dynamic | droplets | Counts the number of droplets in a size class with a mean size of 20 micrometers. |
| *droplets_size28* | Patch, dynamic | droplets | Counts the number of droplets in a size class with a mean size of 28 micrometers. |
| *droplets_size36* | Patch, dynamic | droplets | Counts the number of droplets in a size class with a mean size of 36 micrometers. |
| *droplets_size45* | Patch, dynamic | droplets | Counts the number of droplets in a size class with a mean size of 45 micrometers. |
| *droplets_size62.5* | Patch, dynamic | droplets | Counts the number of droplets in a size class with a mean size of 62.5 micrometers. |
| *droplets_size87.5* | Patch, dynamic | droplets | Counts the number of droplets in a size class with a mean size of 87.5 micrometers. |
| *droplets_size112.5* | Patch, dynamic | droplets | Counts the number of droplets in a size class with a mean size of 112.5 micrometers. |
| *droplets_size137.5* | Patch, dynamic | droplets | Counts the number of droplets in a size class with a mean size of 137.5 micrometers. |
| *droplets_size175* | Patch, dynamic | droplets | Counts the number of droplets in a size class with a mean size of 175 micrometers. |
| *droplets_size225* | Patch, dynamic | droplets | Counts the number of droplets in a size class with a mean size of 225 micrometers. |
| *droplets_size375* | Patch, dynamic | droplets | Counts the number of droplets in a size class with a mean size of 375 micrometers. |
| *droplets_size750* | Patch, dynamic | droplets | Counts the number of droplets in a size class with a mean size of 750 micrometers. |
| *droplets_size3AtInf* | People, static | droplets | Counts the number of droplets in a size class with a mean size of 3 micrometers that were present in the containing patch when the individual was infected. |
| *droplets_size6AtInf* | People, static | droplets | Counts the number of droplets in a size class with a mean size of 6 micrometers that were present in the containing patch when the individual was infected. |
| *droplets_size12AtInf* | People, static | droplets | Counts the number of droplets in a size class with a mean size of 12 micrometers that were present in the containing patch when the individual was infected. |
| *droplets_size20AtInf* | People, static | droplets | Counts the number of droplets in a size class with a mean size of 20 micrometers that were present in the containing patch when the individual was infected. |
| *droplets_size28AtInf* | People, static | droplets | Counts the number of droplets in a size class with a mean size of 28 micrometers that were present in the containing patch when the individual was infected. |
| *droplets_size36AtInf* | People, static | droplets | Counts the number of droplets in a size class with a mean size of 36 micrometers that were present in the containing patch when the individual was infected. |
| *droplets_size45AtInf* | People, static | droplets | Counts the number of droplets in a size class with a mean size of 45 micrometers that were present in the containing patch when the individual was infected. |

| | | | |
|---|---|---|---|
| *droplets_size62.5AtInf* | People, static | droplets | Counts the number of droplets in a size class with a mean size of 62.5 micrometers that were present in the containing patch when the individual was infected. |
| *droplets_size87.5AtInf* | People, static | droplets | Counts the number of droplets in a size class with a mean size of 87.5 micrometers that were present in the containing patch when the individual was infected. |
| *droplets_size112.5AtInf* | People, static | droplets | Counts the number of droplets in a size class with a mean size of 112.5 micrometers that were present in the containing patch when the individual was infected. |
| *droplets_size137.5AtInf* | People, static | droplets | Counts the number of droplets in a size class with a mean size of 137.5 micrometers that were present in the containing patch when the individual was infected. |
| *droplets_size175AtInf* | People, static | droplets | Counts the number of droplets in a size class with a mean size of 175 micrometers that were present in the containing patch when the individual was infected. |
| *droplets_size225AtInf* | People, static | droplets | Counts the number of droplets in a size class with a mean size of 225 micrometers that were present in the containing patch when the individual was infected. |
| *droplets_size375AtInf* | People, static | droplets | Counts the number of droplets in a size class with a mean size of 375 micrometers that were present in the containing patch when the individual was infected. |
| *droplets_size750AtInf* | People, static | droplets | Counts the number of droplets in a size class with a mean size of 750 micrometers that were present in the containing patch when the individual was infected. |
| *dropletDecay* | Global, static | % droplets removed / min | Droplet decay rate. |
| *expectorateHeight* | Global, static | m | The height at which droplets are expelled. This is also the maximum vertical height of the simulated world, and the height used to in area volume calculations. (i.e., patch volumes are 1 m X 1 m X expectorateHeight m). |
| *expectorateRisk* | Person, static | - | Denotes proportion of droplets agents expel on any given timestep. Defaults to 1. Changes if people are wearing masks. |
| *exposureRisk* | Person, static | - | Denotes agents' probability of infection given exposure to infectious agents. Defaults to 1 (i.e., complete susceptibility). |
| *face-northward* | Global, static | - | Logical variable that controls whether people only look northward within a range of 90 degrees (if TRUE) or face a random direction (if FALSE). |
| *firstInfectTime* | Global, dynamic | ticks | Records the tick at which the first transmission event occurs. |
| *grid-height* | Global, static | m | The number of rows present in the grid representing the room in which agents interact. Note: cells in the matrix (i.e., patches), regardless of how many there are, represent 1 m X 1 m areas. |
| *grid-width* | Global, static | m | The number of columns present in the grid representing the room in which agents interact. Note: cells in the matrix (i.e., patches), regardless of how many there are, represent 1 m X 1 m areas. |
| *infected?* | Person, dynamic | - | Logical variable describing if people have been infected by contaminated patches. |
| *infectious?* | Person, static | - | Logical variable describing if people can spread the pathogen. |
| *lastInfectTime* | Global, dynamic | ticks | Records the tick at which the last susceptible individual was infected. Only relevant if ALL susceptible people were infected. |
| *mod_group* | Global, static | - | Controls what agent variables are modified by risk mod. Takes the values "sus," "inf," or "sus_inf" (representing susceptible agents only, infectious agents only, or both). If "sus," only exposureRisk is adjusted. If "inf," only expectorateRisk is adjusted. If "sus_inf," both of these variables are updated. |
| *mod-proportion* | Global, static | - | Describes the probability that susceptible individuals will have their infection probability modified by the maskRisk-mod parameter. This parameter is used to vary the proportion of individuals minimizing their disease risk in the population (e.g., through the use of PPE). |
| *n* | Global, static | people | The total number of (i.e., both "healthy" and "infectious") people turtles that spawn in each cohort. |
| *n_infectious* | Global, static | people | The number of infectious people turtles that spawn in each cohort. This is a subset of n, not additional turtles. Infectious people default to "asymptomatic" status. |
| *num_asymptomatic* | Global, dynamic | people | The number of asymptomatic "infectious" people that spawn in the first cohort. The probability of an infectious agent being asymptomatic is (*n_infectious* *(1 - *symp-pr)).* |

| | | | |
|---|---|---|---|
| *num_symptomatic* | Global, dynamic | people | The number of symptomatic "infectious" people that spawn in the first cohort. The probability of an infectious agent being symptomatic is (*n_infectious* * *symp-pr*). |
| *num_completelySusceptible* | Global, dynamic | people | Counts the number of individuals in a cohort that are completely susceptible to infection. |
| *num_reducedSusceptible* | Global, dynamic | people | Counts the number of individuals in a cohort with reduced susceptibility to infection. |
| *numCohorts* | Global, static | cohorts | The number of people cohorts observed during the simulation. Note: all cohorts interact with the same grid (i.e., world), but do not exist in the world at the same time. |
| *numReturnVents* | Global, static | vents | The number of patches designated as return vents. Cannot exceed grid-width value if ventilReturnWall is one of "north," "south," "up," or "down." Cannot exceed grid-height value if ventilReturnWall is one of "east," "west," "left," or "right." |
| *numSupplyVents* | Global, static | vents | The number of patches designated as supply vents. Cannot exceed grid-width value if ventilSupplyWall is one of "north," "south," "up," or "down." Cannot exceed grid-height value if ventilReturnWall is one of "east," "west," "left," or "right." |
| *patchContamination.list* | Global, dynamic | - | List of the number of contaminated patches present at each time step. |
| *patchInfectiousness.list* | Global, dynamic | - | List of the mean patch infectiousness values observed throughout the simulation. |
| *person-count* | Patch, dynamic | people | Counts the number of people in the cell. |
| *personDist.list* | Global, dynamic | - | List of average distance between people at each tick. |
| *personPerPatch-cap* | Global, static | people | The maximum number of people that may possibly exist within a single patch. |
| *mask?* | Person, static | - | Logical variable describing if people are wearing a mask. |
| *maskRisk-mod* | Global, static | - | The probability that people exposed to virions will be infected after spending 1-tick duration in a contaminated patch when a mask, or that infectious individuals expectorate infectious droplets when wearing the same masks. |
| *rearrange-cohort* | Global, static | - | Logical variable describing whether or not the cohort-replace effectively becomes a rough proxy for movement of people within the room. If TRUE, the first "cohort" never leaves the room, rather, they are re-distributed according to the social-distance and personPerPatch-cap values set. |
| *returnVent* | Patch, static | - | Denotes if patch is a return vent. |
| *showArrows* | Global, static | - | Logical variable controling if airArrow turtles will be hidden or not. If TRUE, airArrows will be visible. If FALSE, they will be hidden. |
| *social-distance* | Global, static | m | The interpersonal distance that people seek to maintain over the course of the simulation. |
| *speak_airflow-angle* | Global, static | degrees | The angle of airflow associated with breathing events. Affects the spread of droplets during a given droplet-expulsion event from asymptomatic individuals. |
| *speak_spread-dist.mean* | Global, static | m | The mean distance from asymptomatic infectious people that droplets may be spread when breathing. This value will be used to generate a lognormal distribution from which droplet-expulsion-spread distance will be randomly drawn when an asymptomatic agent triggers a droplet-expulsion event. |
| *speak_spread-dist.sd* | Global, static | m | The standard deviation distance, given an speak_spread-dist.mean value, from asymptomatic infectious people that droplets may be spread when breathing. This value will be used to generate a lognormal distribution from which droplet-expulsion-spread distance will be randomly drawn when an asymptomatic agent triggers a droplet-expulsion event. |
| *supplyVent* | Patch, static | - | Denotes if patch is a supply vent. |
| *symp-pr* | Global, static | - | Probability that infectious people will be "symptomatic," instead of having the default "asymptomatic" status. |
| *symptomatic?* | Person, static | - | Logical variable describing if people are coughing to spread the contagion. If TRUE, spread will be dictated by cough_airflow-angle, cough_spread-dist.mean, and cough_spread-dist.sd parameter values. If FALSE, but infectius? is TRUE, spread will be dictated by speak_airflow-angle, speak_spread-dist.mean, and speak_spread-dist.sd parameter values. |
| *totalDroplets* | Patch, dynamic | droplets | Counts the total number of droplets present in the patch. |

| | | | |
|---|---|---|---|
| *totalInfected* | Global, dynamic | people | Running sum of the total number of infected people over the course of the simulation. |
| *totalInfected.list* | Global, dynamic | - | List of the total number of infected people at each tick. |
| *transmissionRisk* | Patch, dynamic | - | Tracks the probability that susceptible people on the patch will be infected on a given time point. This is the product of virionCount and virionRisk. |
| *ventilation* | Global, static | - | Logical variable describing if airflow will move droplets throughout patches during the simulation. |
| *ventil_movementRate* | Global, static | % air change / min | Describes the rate at which air (and therefore droplets suspended in the air) will move to another patch at each tick if ventilation effects are being simulated. |
| *ventil_removalRate* | Global, static | - | Describes the proportion of droplets on return vent patch(es) that will be removed from the simulation due to filtration. |
| *ventilReturnWall* | Global, static | - | Takes one value "north," "south," "east," "west," OR "up," "down," "right," "left." Describes the wall of the simulated world that return vents will be located on. |
| *ventilSupplyWall* | Global, static | - | Takes one value "north," "south," "east," "west," OR "up," "down," "right," "left." Describes the wall of the simulated world that supply vents will be located on. |
| *virionCount* | Patch, dynamic | virions | Counts the number of virions in the patch on a given time step. |
| *virionRisk* | Global, static | - | The risk of infection given exposure to a single virion. |
| *virionsPerML* | Global, static | virions / mL | Number of virions per mL of droplet fluid. |
| *vol_B* | Global, static | $m^3$ / min | The rate of air inhaled by individuals in patches. |
| *Vt_diam3* | Global, static | m / min | The terminal velocity of a respiratory droplet with a 3-micrometer diameter, calculated from equations given by Anchordopqui & Chudnovsky (2020). |
| *Vt_diam6* | Global, static | m / min | The terminal velocity of a respiratory droplet with a 6-micrometer diameter, calculated from equations given by Anchordopqui & Chudnovsky (2020). |
| *Vt_diam12* | Global, static | m / min | The terminal velocity of a respiratory droplet with a 12-micrometer diameter, calculated from equations given by Anchordopqui & Chudnovsky (2020). |
| *Vt_diam20* | Global, static | m / min | The terminal velocity of a respiratory droplet with a 20-micrometer diameter, calculated from equations given by Anchordopqui & Chudnovsky (2020). |
| *Vt_diam28* | Global, static | m / min | The terminal velocity of a respiratory droplet with a 28-micrometer diameter, calculated from equations given by Anchordopqui & Chudnovsky (2020). |
| *Vt_diam36* | Global, static | m / min | The terminal velocity of a respiratory droplet with a 36-micrometer diameter, calculated from equations given by Anchordopqui & Chudnovsky (2020). |
| *Vt_diam45* | Global, static | m / min | The terminal velocity of a respiratory droplet with a 45-micrometer diameter, calculated from equations given by Anchordopqui & Chudnovsky (2020). |
| *Vt_diam62.5* | Global, static | m / min | The terminal velocity of a respiratory droplet with a 62.5-micrometer diameter, calculated from equations given by Anchordopqui & Chudnovsky (2020). |
| *Vt_diam87.5* | Global, static | m / min | The terminal velocity of a respiratory droplet with a 87.5-micrometer diameter, calculated from equations given by Anchordopqui & Chudnovsky (2020). |
| *Vt_diam112.5* | Global, static | m / min | The terminal velocity of a respiratory droplet with a 112.5-micrometer diameter, calculated from equations given by Anchordopqui & Chudnovsky (2020). |
| *Vt_diam137.5* | Global, static | m / min | The terminal velocity of a respiratory droplet with a 137.5-micrometer diameter, calculated from equations given by Anchordopqui & Chudnovsky (2020). |
| *Vt_diam175* | Global, static | m / min | The terminal velocity of a respiratory droplet with a 175-micrometer diameter, calculated from equations given by Anchordopqui & Chudnovsky (2020). |
| *Vt_diam225* | Global, static | m / min | The terminal velocity of a respiratory droplet with a 225-micrometer diameter, calculated from equations given by Anchordopqui & Chudnovsky (2020). |
| *Vt_diam375* | Global, static | m / min | The terminal velocity of a respiratory droplet with a 375-micrometer diameter, calculated from equations given by Anchordopqui & Chudnovsky (2020). |
| *Vt_diam750* | Global, static | m / min | The terminal velocity of a respiratory droplet with a 750-micrometer diameter, calculated from equations given by Anchordopqui & Chudnovsky (2020). |

### 10. *Figures*

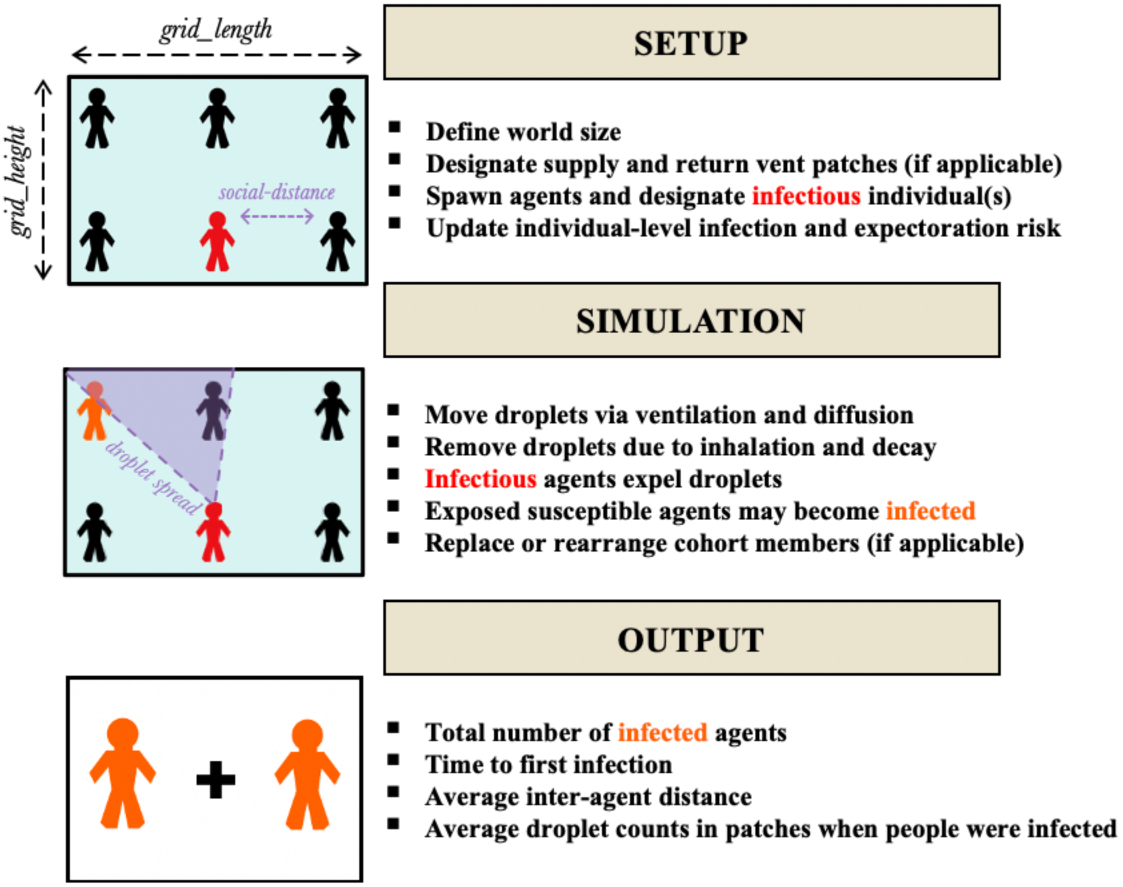


Figure S1-1. Simplified model overview. Bulleted sub-models are listed in the order that they take place within the model. Simulation bullet points repeat each model tick.

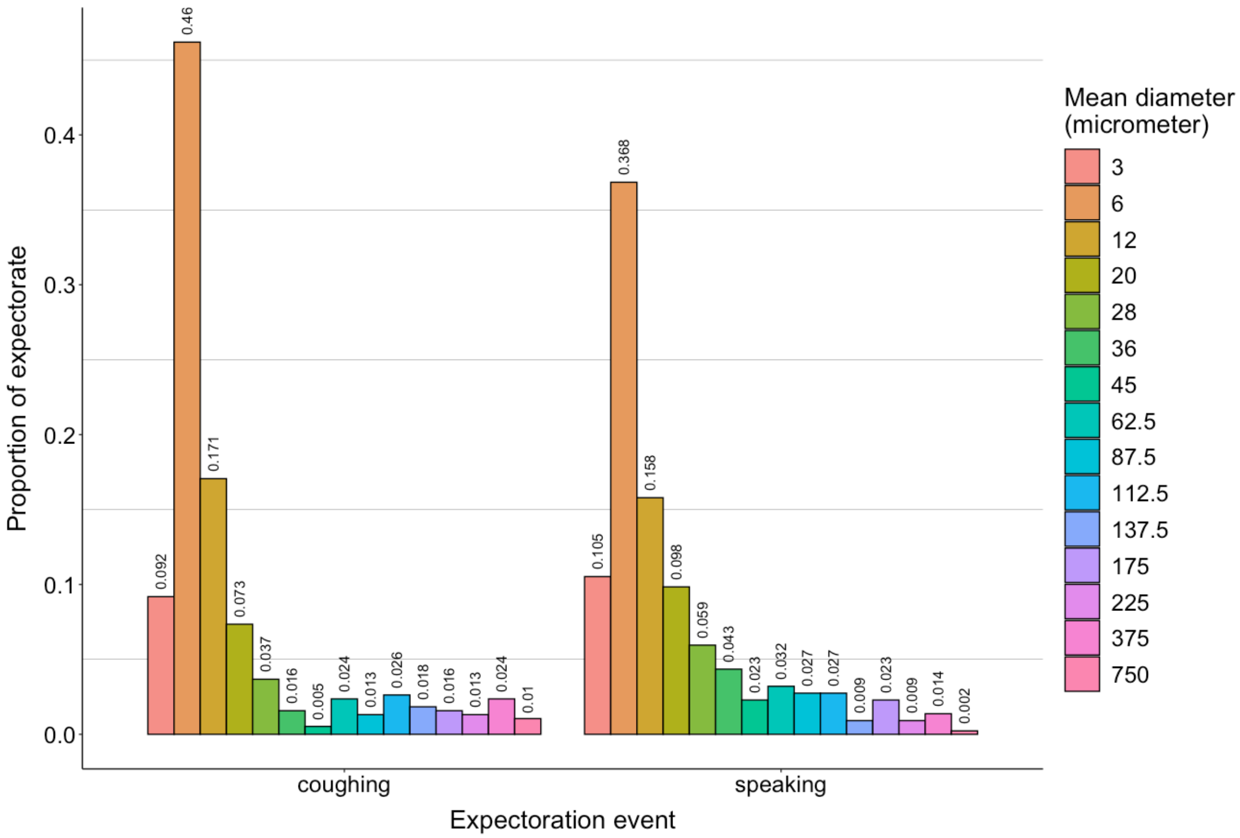


Figure S1-2. Distribution of droplet sizes during expectoration events. Distributions of size classes during coughing and speaking events are based on findings of Chao *et al.* (2009), and represent mean observed droplet-size measurements they recorded 60 mm away from individuals' mouths immediately following these activities.

a.)

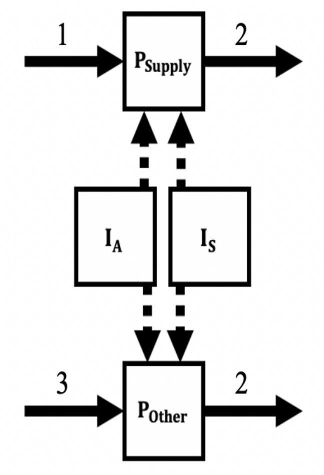


b.)

| Number | Equation | Description |
|---|---|---|
| 1 | $\frac{\left(\sum_{r=1}^{P_{Return}}\left(num_{d_r} * ventil\_movementRate * (1 - ventil\_removalRate)\right)\right)}{P_{Supply}} + \sum_{n=1}^{Neighbors}\left(\sum_{d=1}^{D}\left(num_{d_n} * (ventil\_movementRate + diffusionRate)\right)\right)$ | Fixed droplet input to supply patches |
| 2 | $\sum_{d=1}^{D}\left(\left(\left(\left(\frac{vol_B}{1\,\mathrm{m} * 1\,\mathrm{m} * expectorateHeight} * People_p\right) + \left(\frac{Vt_{diam_d}}{expectorateHeight}\right) + dropletDecay\right) + ventil\_movementRate + diffusionRate\right) * num_{d_p}\right)$ | Droplet removal from patches |
| 3 | $\sum_{n=1}^{Neighbors}\left(\sum_{d=1}^{D}\left(num_{d_n} * (ventil\_movementRate + diffusionRate)\right)\right)$ | Fixed droplet input to non-supply patches |

Figure S1-3. Droplet dynamics for supply-vent and non-supply-vent patches. a.) When modeling ventilation, droplet input to and removal from patches are functions of fixed rates (solid arrows) and probabilistic expectoration from symptomatic and asymptomatic people within range (dashed arrows). b.) Equations for fixed effects on within-patch droplet dynamics. Supply patches receive input from Return-vent patches in addition to diffusion from nearby neighbors. Non-supply-vent patches do not receive input from Return vents. Droplet output is a function of ventilation airflow parameters, diffusion to neighbors, a fixed decay rate, and inhalation by people within the patch.
