## Appendix S2 for "Assessing the efficacy of interventions to control indoor SARS-Cov-2 transmission: an agent-based modeling approach"

### Appendix S2 – Rationale for model parameterization to simulate a SARS-CoV-2 superspreading event

As noted in the main text, for benchmarking purposes, we simulated the Skagit County, Washington, USA March 2020 SARS-CoV-2 superspreading event as case scenario. Because this superspreading event is thought to be the result of transmission from a single infectious individual (Hamner *et al.* 2020), all simulations contained only one infectious person. The infectious person was assumed to be symptomatic during the choir practice. We assumed all droplets were expelled from this individual at a height of 1.7 m, the approximate mean height of U.S. adults (Fryar *et al.* 2018). We make the assumption that the cough frequency for a symptomatic COVID-19 patient is equal to that of individuals with a chronic cough condition. Therefore, every minute our infectious individual had a 19% probability to expel droplets through coughing (Lee *et al.* 2012), and an 81% chance to expel droplets through an unspecified other activity (e.g., speaking, singing, etc.). Using the procedure described by Railsback & Grimm (2011), droplet travel distances for coughing and non-coughing expectoration events were randomly drawn from lognormal distributions with known means and standard deviations. Travel distances for coughing events were drawn from a distribution with a mean of 5 m and standard deviation of 0.256 m (Bourouiba *et al.* 2014). Travel distances for non-coughing events were drawn from a distribution with a mean of 0.55 m and standard deviation of 0.068 m (Das *et al.* 2020). The angle of droplet spread during coughing and non-coughing expectorations were $35^{o}$ and $63.5^{o}$, respectively, in accordance with median values of mouth-angle ranges described by Kwon *et al.* (2012). We set the inhalation rate for simulated individuals to 0.023 m3 air/min, a rate consistent with adults participating in light activity (Adams 1993).

We know from the Hamner et al. (2020) case report that the choir practice lasted 150 minutes in total, split into 4 distinct time intervals lasting 40, 50, 15, and 45 minutes. During the first time interval, all 61 attendees practiced together in the 180 m2 main hall for 40 minutes. In the second interval, the group split into two subsets of unspecified sizes. One subset rearranged themselves within the main hall, and the second subset moved into a separate room. The subsets rehearsed separately for 50 minutes. The third time interval was a 15-minute break period when individuals mixed freely. During the final time interval, all attendees returned to the main hall to practice as a single group once more for 45 minutes. When practicing as single group during intervals 1 and 4, individuals sat in assigned seats (Miller et al. 2020) with chairs spaced 15.24 – 25.4 cm apart (Hamner et al. 2020). In our simulations, we decided to rearrange agents in our model after 40, 90, and 105 minutes to recreate mixing associated with changing time intervals. At timestep 105, individuals moved back to their initial placements, representing their adherence to assigned seating during interval 4 (i.e., minutes 105 – 150). The seating chart has not been shared due to privacy concerns (Miller et al. 2020) however, from the spacing estimate we can assume that a maximum of 2 people could be within 1-$m^2$ patches in our model scenario. Our ability to simulate mixing rates during specific time intervals was limited to this extent because we do not know specific seating arrangements, subset size or configuration, secondary room size, or interaction rates during the break period.

Miller *et al.* (2020) estimated that the infectious individual in the Skagit County case study emitted 970 ± 390 SARS-CoV-2 quanta/hr. A quantum is the number of aerosolized infectious particles required to infect 1- 1/e % (i.e., ≈ 63%) of a susceptible population, assuming that all individuals were exposed to the same number of particles (Wells 1955). Here we describe how we estimated the virion risk associated with 1 quanta $min^{-1}$ to be used in our SARS-CoV-2 transmission efforts. To do this, we ran a modified version of our agent-based model, wherein the droplet fallout procedure (see Supplemental Materials I) was carried out before the infection procedure, and droplets were homogenously dispersed throughout the entirety of the simulated world immediately after expectoration. We parameterized our modified model to reflect the choral super-spreading event described by Hamner *et al.* 2020 (Table S2-1). We varied 22 virion infection risk levels across 220,000 simulations (i.e., 10,000 simulations per level). All simulations lasted only a single time step, after which we recorded the percentage of susceptible individuals infected. After evaluation, all simulations were aggregated into a single data set, and we carried out a linear regression to relate the percentage of susceptible people infected to the virion risk:

$$\%\text{ infected} = \beta_0 + \beta_1 virionRisk.$$

We determined that

$$\%\text{ infected} = 0.0027 + 10.06 virionRisk.$$

Given this formula, we calculated that a virion risk value of 0.0624 is required to infect 63% of susceptible in our parameterized quantum-simulation model. We adopted this value as the virion risk in all primary simulations.

The number of droplets produced by the infectious individual in our simulations each minute was independent of coughing status, and like droplet travel distance estimation, was drawn from a log-normal distribution. The known mean and standard deviation values for this distribution were $9.7e^5$ and $3.9e^5$ droplets, respectively, to recreate the 970 ± 390 quanta/hr estimated by Miller *et al.* (2020). We assumed a virion decay rate of 1.05 % /min (van Doremalen *et al.* 2020).

We assume that airborne droplets naturally diffuse throughout the simulated environment at a fixed rate of $1.5e^{-3}$ $m^3$/min (Castillo & Weibel 2018) regardless of size. Additionally, we know that the ventilation system in the main hall of the church consists of three supply vents that push a mixture of outdoor and recirculated air towards a single return vent on the opposite wall, though the true direction of forced airflow (e.g., North to South) is unclear from reports (Miller et al. 2020). Because it is uncertain whether or not the forced-air system was turned on during the choir practice (Miller et al. 2020), however, we decided to run our simulations in two sets: ventilation-on (i.e., both forced-air effects and natural diffusion moved droplets between patches) and ventilation-off (i.e., only natural diffusion moved droplets between patches). In the ventilation-on set, we additionally assume that droplets move from supply vents towards the return vent at a fixed rate of 0.043 %/min (Miller *et al.* 2020), and that 90% of droplets were filtered prior to recirculation (Miller *et al.* 2020). Because we do not know the true direction of forced airflow, we simulated both North-to-South and East-to-West forced airflow movement in the ventilation-on set (Figure S2-1).

*References*

*Tables*

Table S2-1. Parameters for SARS-CoV-2 quantum simulation.

| Parameter | Value(s) | Reference(s) |
|---|---|---|
| Area ($m^2$) | 180 | Hamner *et al.* 2020 |
| Cough frequency (coughs/min) | 0.19 | Lee *et al.* 2012 |
| Droplet count (droplets/expectoration) | 60,000 | Stadnytskyi *et al.* 2020 |
| Expectoration height (m) | 1.7 | Fryar *et al.* 2018 |
| Inhalation rate ($m^3$ air/min) | 0.023 | Adams 1993 |
| Maximum people in a single 1-$m^2$ patch (people) | 2 | Hamner *et al.* 2020 |
| virion count (virions/mL fluid) | $2.35e^9$ | Wölfel *et al.* 2020 |
| virion decay rate (%/min) | 1.05 | van Doremalen *et al.* 2020 |
| virion infection risk (%/inhaled virion) | $1.0e^{-5}$, $3.0e^{-5}$, $5.0e^{-5}$, $8.0e^{-5}$, $1.0e^{-4}$, $3.0e^{-4}$, $5.0e^{-4}$, $8.0e^{-4}$, 0.001, 0.003, 0.005, 0.008, 0.01, 0.02, 0.03, 0.04, 0.05, 0.06, 0.07, 0.08, 0.09, 0.1 | – |

*Figures*

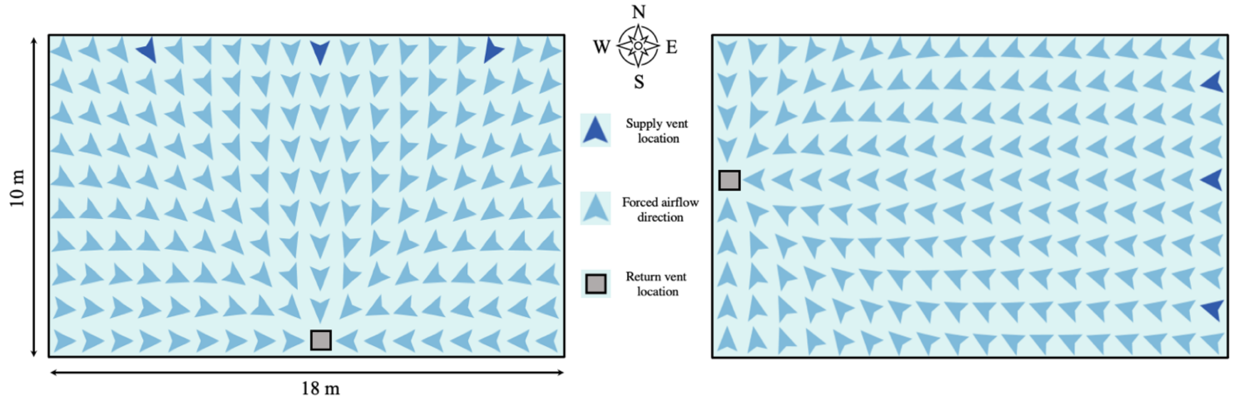

Figure S2-1. Airborne infectious droplets in North-to-South and East-to-West forced airflow schemas have different maximum travel distances due to the shape of the simulated world.
