## Appendix S3 for "Assessing the efficacy of interventions to control indoor SARS-Cov-2 transmission: an agent-based modeling approach"

### Appendix S3 – Sensitivity analysis

We carried out a global sensitivity analysis to estimate Sobol' indices for select parameters using formulas given by Jansen (1999) and Saltelli *et al.* (2010), commonly referred to as "Jansen estimators." This method allows us to simultaneously estimate 1st order and total Sobol' indices, estimates of the model variance attributed to effects of the parameter alone and attributable to the singular parameter effect plus all interactions with other varied parameters, respectively. Due the size of our model's full parameter space and computational limitations, we included only infectiousness parameters in the sensitivity analysis (Table S3-1). The included parameters were chosen because their values are obligate for any study aimed at estimating infection rates using our model. Airflow and most intervention parameters were excluded. These parameters are situation specific, and may be varied in case scenarios to test specific hypotheses.

The number of model evaluations required when using Jansen estimators is equivalent to $(k + 2)*S$, where $k$ is the number of parameters included in the analysis, and $S$ is the number of random samples drawn from parameter ranges (Saltelli *et al.* 2010). Based on 1250 random samples, we ran 25,000 simulations of our model across three random seeds (Salecker *et al.* 2019), therefore running 75,000 simulations in total. We used 5,000 bootstrap replicates to generate estimates for each random seed level. We report estimated Sobol' indices averaged across random seed levels in Table S3-2. This sensitivity analysis was carried out using the NLRX R package (Salecker *et al.* 2019) in RStudio (v. 1.1.463, RStudio Team, Boston, MA) running R (v. 3.6.2, R Foundation for Statistical Computing, Vienna, Austria).

*References*

*Tables*

Table S3-1. Values used to parameterize simulations for sensitivity analysis.

| Parameter | Range |
|---|---|
| varied parameters | |
| *cohort-dur* | 2 – 90 |
| *cough_airflow-angle* | 35 – 90 |
| *cough_dropletNum.mean** | $1e^{4}$ – $1e^{5}$ |
| *cough_dropletSizeDistr*§ | 1 – 4 |
| *cough_frequency* | 0 – 0.25 |
| *cough_spread-dist.mean** | 2 – 7 |
| *diffusionRate* | 0.001 – 0.01 |
| *dropletDecay* | 0.01 – 0.25 |
| *expectorate-height* | 0.5 – 2 |
| *grid-height* | 3 – 10 |
| *grid-width* | 3 – 10 |
| *speak_airflow-angle* | 35 – 90 |
| *speak_dropletNum.mean** | $1e^{4}$ – $1e^{5}$ |
| *speak_dropletSizeDistr*§ | 1 – 4 |
| *speak_spread-dist.mean** | 0.5 – 2 |
| *virionRisk* | $1e^{-6}$ – 0.001 |
| *virionsPerML* | $1e^{7}$ – $1e^{8}$ |
| *vol_B* | 0.01 – 0.025 |
| fixed parameters | |
| *face-northward* | false |
| *mod-proportion* | 0† |

| | |
|---|---|
| *n* | 2 |
| *n_infectious* | 1 |
| *num-cohorts* | 1 |
| *personPerPatch-cap* | 2 |
| *rearrange-cohort* | false |
| *social-distance* | 0 |
| *symp-pr* | 1 |
| *ventilation* | false[‡] |

[*]All standard deviation parameters associated with “mean” parameters were set to 0. [†]Because *mod-proportion* is 0, *maskRisk-mod* and *mod-group* values are irrelevant. [‡]Because *ventilation* is false, all ventilation-related parameter values (e.g., *ventil_movementRate, numSupplyVents, numReturnVents*, etc.) are irrelevant. [§]We were interested in estimating effects of droplet sizes on infection probability. To do so, rather than using droplet size distributions described by Chao *et al.* (2009) to parameterize expectorate, we used the *dropletSizeDistr* parameters. The *dropletSizeDistr* parameter describes the log mean of lognormal distribution with log SD of 0.3 used to define the probability distribution for droplet sizes.

Table S3-2. Mean 1st-order and total Sobol’ indices for infectiousness parameters. 95% confidence intervals are given in parentheses. Parameters are listed in order of descending total Sobol’ index.

| **Parameter** | **1st-Order** | **Total** |
|---|---|---|
| *grid-width* | 0.094 (0.00, 0.308) | 0.873 (0.723, 0.947) |
| *grid-height* | 0.116 (0.017, 0.320) | 0.814 (0.687, 0.918) |
| *cough_airflow-angle* | 0.070 (0.014, 0.226) | 0.183 (0.116, 0.239) |
| *cohort-dur* | 0.059 (0.016, 0.219) | 0.172 (0.108, 0.227) |
| *cough_spread-dist.mean* | 0.075 (0.017, 0.232) | 0.172 (0.107, 0.227) |
| *cough_frequency* | 0.104 (0.013, 0.255) | 0.158 (0.098, 0.209) |
| *cough_dropletSize* | 0.065 (0.010, 0.219) | 0.149 (0.090, 0.200) |
| *virionRisk* | 0.091 (0.021, 0.240) | 0.136 (0.078, 0.184) |
| *speak_spread-dist.mean* | 0.096 (0.024, 0.249) | 0.131 (0.078, 0.177) |
| *virionsPerML* | 0.082 (0.012, 0.232) | 0.118 (0.065, 0.165) |
| *speak_dropletSizeDistr* | 0.071 (0.014, 0.215) | 0.058 (0.021, 0.090) |
| *cough_dropletNum.mean* | 0.087 (0.005, 0.231) | 0.055 (0.020, 0.085) |
| *expectorate-height* | 0.065 (0.013, 0.209) | 0.054 (0.018, 0.084) |

| | | |
|---|---|---|
| *vol_B* | 0.078 (0.012, 0.218) | 0.054 (0.017, 0.084) |
| *dropletDecay* | 0.075 (0.011, 0.220) | 0.052 (0.015, 0.081) |
| *speak_airflow-angle* | 0.088 (0.018, 0.231) | 0.046 (0.014, 0.073) |
| *diffusionRate* | 0.068 (0.008, 0.210) | 0.042 (0.01, 0.068) |
| *speak_dropletNum.mean* | 0.075 (0.007, 0.217) | 0.039 (0.010. 0.064) |
